# Agentic AI for automated hypothesis testing in Alzheimer’s disease and related dementias

**DOI:** 10.64898/2025.12.02.25341517

**Authors:** Ketan S. Saichandran, Karim Elzokm, Osman B. Guney, Vijaya B. Kolachalama

**Affiliations:** Department of Medicine, Boston University Chobanian & Avedisian School of Medicine, Boston, MA, USA – 02118; Department of Computer Science, Boston University, Boston, MA, USA – 02215; Department of Electrical & Computer Engineering, Boston University, Boston, MA, USA – 02215; Faculty of Computing & Data Sciences, Boston University, Boston, MA, USA– 02215

**Author notes:** Corresponding author, Address: 72 E. Concord St, Evans 636, Boston, MA – 02118.

**Keywords:** Artificial intelligence, Agentic AI, Hypothesis testing

## Abstract

**Background:** Alzheimer’s disease and related dementias (ADRD) are multifactorial disorders driven by genetic, vascular, inflammatory, lifestyle, and biological factors interacting over decades, requiring population-level analyses. However, utilizing multimodal datasets is labor-intensive, impeding discovery and equity. Multi-agentic AI systems promise workflow automation but lack ADRD-specific adaptations.

**Methods:** We created a modular AI system with agents for data preparation, planning, analysis, critique, and summarization. Using data from the National Alzheimer’s Coordinating Center (NACC) (n=48,876), we tested 100 literature-derived ADRD hypotheses, subsampled cohorts (100–10,000 participants), and 100 non-ADRD controls via iterative loops.

**Results:** The system verified 42.0±2.8 hypotheses by rejecting the null, found no evidence for 16.6±1.1, and deemed 41.4±2.3 not testable, with consistency in negated versions. Verification rates rose from 10.4 (n=100) to 35.4 (n=10,000); controls showed 85.2±0.6 non-testable.

**Conclusions:** Our system illustrates a powerful paradigm in which AI can facilitate automated data analysis, expediting advancements in population-level dementia investigations.

## 1 BACKGROUND

Alzheimer’s disease (AD) and related dementias (ADRD) arise from multifactorial causes that include genetic susceptibility, vascular and inflammatory processes, lifestyle influences, and other biological pathways interacting over years.^1^ Understanding how these factors intersect to shape disease onset and progression demands population-level analyses. Typically, these analyses are performed on datasets containing tens to thousands of variables ranging from demographics, comorbidities, and neuropsychological assessments to neuroimaging, fluid biomarkers, and genetics, reflecting the layered nature of ADRD pathophysiology. A wealth of population-based cohorts such as the Alzheimer’s Disease Neuroimaging Initiative,^2^ and the National Alzheimer’s Coordinating Center (NACC),^3, 4^ among others, offer unparalleled access to rich, multimodal data resources. These datasets enable exploration of complex associations such as amyloid-vascular interactions,^5, 6^ or ethnicity-specific trajectories,^7–9^ but leveraging them remains laborious and fragmented.^10^ Researchers often spend significant time to identify variables, align data dictionaries, harmonize features and manage missingness, even for seemingly simple hypotheses (e.g., whether APOE4 modulates vascular effects on hippocampal atrophy).^11^ This friction slows discovery, increases analytic errors, and widens disparities between well-resourced and smaller research teams.

Recent years have witnessed efforts to standardize multimodal ADRD research through coordinated data infrastructures, analytic tools and AI-driven multimodal analysis.^12–14^ Platforms such as the National Institute on Aging Genetics of Alzheimer’s Disease Data Storage Site Data Sharing Service,^15, 16^ and the AD Workbench,^17^ have improved accessibility and interoperability of large-scale cohorts. Technical standards such as the Brain Imaging Data Structure,^18^ and Findable, Accessible, Interoperable, Reusable (FAIR) data principles,^19^ have enabled more consistent data exchange across repositories. Machine learning frameworks like MONAI,^20^ have been developed to analyze neuroimaging data, while federated learning and harmonization methods have addressed privacy-preserving analysis across sites.^21^ Despite these advances, most systems remain specialized for discrete tasks, such as image harmonization, and do not integrate the full scientific workflow from hypothesis generation to testing. In parallel, recent developments in multi-agentic AI systems have begun transforming how scientific discovery can be automated. These systems, which include specialized agents each responsible for tasks like hypothesis formulation, experimental design, or critique, can collaborate autonomously to conduct literature reviews, run simulations, and refine hypotheses based on results.^22–24^ They showcase the feasibility of embedding scientific reasoning within collaborative AI architectures, reducing manual burden while maintaining interpretability and rigor. Yet, most of them operate in general-purpose or simulated environments, lacking adaptation to the specific data standards, ontologies, and multimodal complexities of domains such as ADRD. What remains missing is a unifying, domain-aware and modular system that can autonomously handle data, translate unstructured research queries into analytic hypotheses, and test them rigorously. Such a system would bring conceptual automation to discovery itself, bridging data analysis and hypothesis testing in an iterative, reproducible, and extensible manner.

Here, we developed a multi-agentic AI system for accelerated hypothesis testing in ADRD research. This system combines modular reasoning with robust data processing, allowing each component to be replaced or improved without disrupting the overall workflow. The system comprises specialized agents, which collaboratively process spreadsheets representing cohort-level data and their corresponding data dictionaries. The workflow operates through two nested loops: an outer loop where the data preparation and planning agents filter relevant variables and outline analysis plans, and an inner loop where the scientist and critic agents execute statistical analyses and perform bias or reproducibility checks. User inputs, ranging from natural language hypotheses to folders containing raw data, are converted into structured analytic pipelines. The critic agent determines outcomes, followed by generation of plots and summaries. We demonstrated the system’s robustness across multiple scenarios: automated testing and replication of literature-derived ADRD hypotheses on the NACC cohort, verifying known associations and identifying non-testable claims; stable performance across dataset sizes; and high specificity in a negative control using randomly selected non-ADRD hypotheses, where majority were correctly classified as not testable on NACC data. As proof of concept, our system points toward a new paradigm, where AI systems can automatically analyze data and accelerate progress in population-level dementia research.

## 2 METHODS

### 2.1 Study population

The study utilized data from the National Alzheimer’s Coordinating Center (NACC), which aggregates information from multiple cohorts and is based on the Uniform Data Set (UDS) 3.0 dictionary.^25^ A total of 48,876 participants were included, with eligibility based on a diagnosis of normal cognition (NC), mild cognitive impairment (MCI), or dementia. We did not exclude cases based on the absence of features or diagnostic labels. For individuals from the NACC cohort with multiple clinical visits, we prioritized visits at which the participant received a dementia diagnosis; among these, we selected the visit with the most available data features, prioritizing neuroimaging information, and chose the most recent visit in cases of ties. This approach maximized the sample sizes of dementia cases, ensured inclusion of each individual’s latest record, and optimized the utilization of available data. Various features (n=750) were included, encompassing subject demographics, medical history, laboratory results, medications, neuropsychological tests, and functional assessments. All participants or their designated informants provided written informed consent.

### 2.2 Hypotheses curation

To curate the literature-derived hypotheses for evaluating agentic AI’s performance, we first compiled a bag of keywords encompassing key ADRD domains, such as “*Alzheimer’s disease risk factors*,” “*dementia biomarkers*,” “*APOE4 interactions*,” “*vascular contributions to neurodegeneration*,” “*ethnicity-specific AD trajectories*,” and related terms drawn from established literature in the field. Using a large language model (ChatGPT, based on GPT-4), we prompted it to generate hundreds of diverse hypotheses inspired by these keywords, along with suggested peer-reviewed references supporting each one. We then manually reviewed each generated hypothesis for scientific plausibility and selected 100 hypotheses relevance to population-level analyses, and alignment with variables available in the NACC cohort; for each, we accessed the original publications (primarily from 2000 to 2024) to verify accuracy, extract supporting excerpts or quotes, and resolve any discrepancies or inaccuracies through team consensus. Hypotheses were refined for clarity, feasibility within our system, and diversity across pathophysiological pathways, resulting in a final set that represented a broad spectrum of testable ADRD research questions while excluding those requiring unavailable data modalities. The list of hypotheses along with corresponding references are provided in **Supplementary Table 1**.

### 2.3 Multi agentic system

Our agentic system employs four specialized large language model (LLM) agents, data preparation agent, planning agent, scientist agent, and critic agent, which collaborate to conduct comprehensive analyses. The pipeline begins with the data preparation agent, where relevant variables are automatically selected based on hypothesis relevance. Following data preparation, the planning agent designs the analytical strategy and recommends appropriate statistical tests and methodologies. Subsequently, an inner loop cycle is initiated between the scientist and critic agents. The scientist agent, following the plan, writes and executes code to conduct required analyses. The critic agent evaluates the results, either approving or rejecting the analysis, and provides feedback to the scientist agent. This inner loop continues until either the hypothesis is approved (i.e., null rejected), rejected (i.e., null not rejected), or determined to be not testable. If not approved, the system may return to data preprocessing with alternative variable selections in an outer loop iteration, allowing for refinement and exploration of different analytical approaches. The system overview is presented in **Figure 1**. Detailed descriptions of each agent are provided below and prompts used on the agentic system are provided in **Supplementary Table 2**.

**Figure 1:**
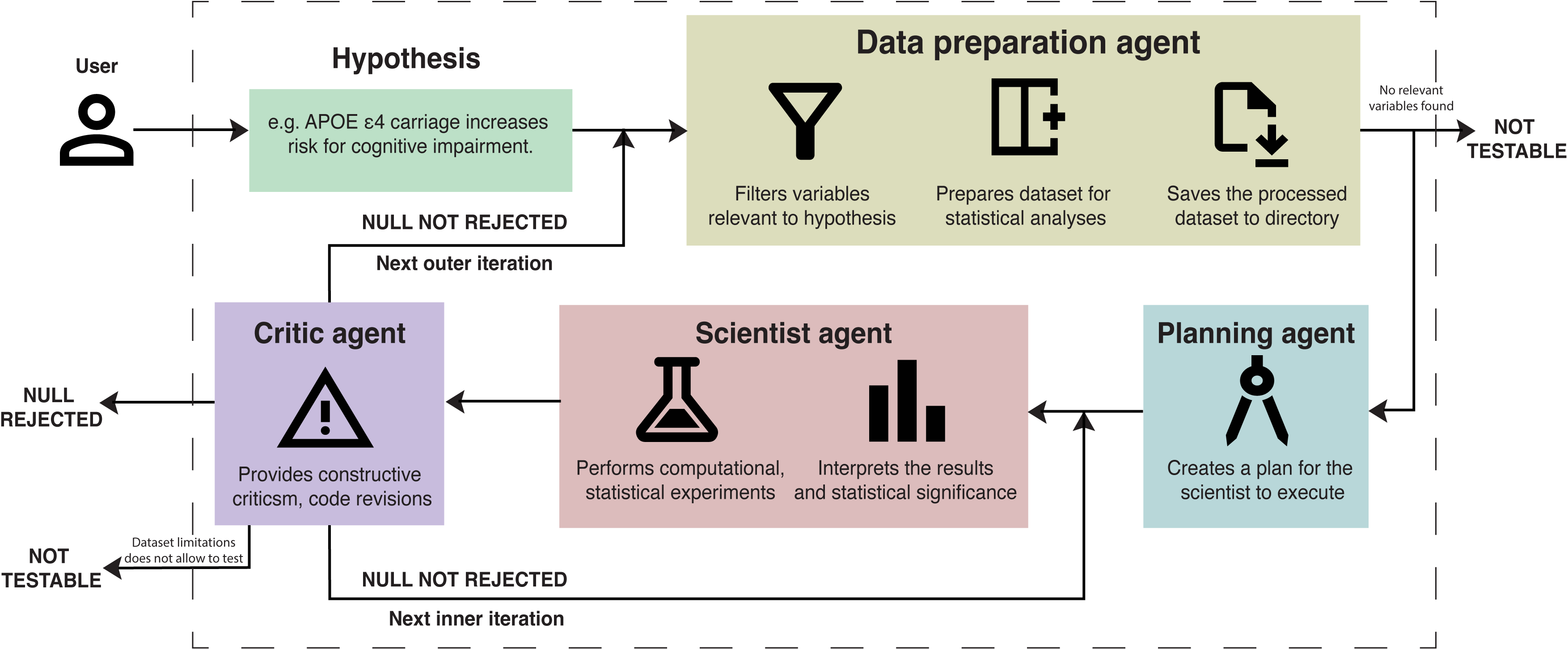
Agentic AI for automated hypothesis testing. The diagram depicts a multi-agent iterative workflow for evaluating user-provided hypotheses (e.g., “*APOE* ε*4 carriage increases risk for Alzheimer’s disease across ethnicities*”). The process begins with the Data Preparation Agent, which filters relevant variables, prepares datasets for analysis, and saves processed data. If no relevant variables are found, the hypothesis is deemed “NOT TESTABLE.” Otherwise, if the null hypothesis is not rejected, the system proceeds to an outer iteration. A core inner loop involves: the Critic Agent (providing constructive criticism and code revisions), the Scientist Agent (performing computational/statistical experiments, interpreting results, and assessing significance), and the Planning Agent (creating execution plans). Additional “NOT TESTABLE” paths account for dataset limitations. The system supports collaboration with external resources (e.g., PubMed) and ADRD data inputs (cohort metadata, CSVs), enabling use cases such as replication of known associations, cross-cohort hypothesis testing, and generation of ranked, interpretable outputs.

#### Data preparation agent

The data preparation agent automatically detects tabular data files (CSV format) and their corresponding data dictionaries (in JavaScript Object Notation (JSON) format) within the workspace folder. It loads both into a centralized cache that stores the dataset along with rich metadata, including variable names, descriptions, inferred data types, and pre-computed statistical summaries. For each user-provided hypothesis, the agent uses the hypothesis text together with the full variable dictionary to automatically select the most relevant features.

Selected variables are then systematically preprocessed: cohort-specific missing-value codes are converted to NaN, categorical variables are one-hot encoded, and continuous/integer variables are standardized as needed. The cleaned, processed dataset is saved to the working directory, and both its file path and updated variable descriptions are injected into the conversation context for downstream agents.

#### Planning agent

The planning agent designs multi-step research strategies and analytical systems, providing methodological recommendations and validation before analysis begins. After adding the generated plan to the conversation context, the system enters an iterative loop between the scientist agent and the critic agent, which we term as the “inner loop”.

#### Scientist agent

The scientist agent performs comprehensive statistical analysis, including exploratory data analysis (EDA), hypothesis testing, regression modeling (linear, logistic, survival), machine learning approaches, mediation and moderation analysis, and subgroup analyses. Before each analysis, the system automatically provides the scientist agent with detailed variable descriptions, data types, and statistical summaries, ensuring informed analytical decisions. The agent then generates Python code for statistical computations and executes analyses with error handling and missing data management. The code is run in a packaged and secure environment to ensure safety and prevent unintended system modifications.

#### Critic agent

The critic agent reviews and validates scientific approaches, providing methodological feedback, and determining hypothesis verification status. The agent evaluates code correctness, statistical validity, result interpretation, and hypothesis alignment, ensuring rigorous scientific standards are maintained. The critic is guided by a comprehensive prompt that instructs it to examine multiple aspects of the analysis: reasonable statistical analysis and adequate sample sizes, sound experimental design that properly tests the hypothesis, appropriate data preprocessing and feature engineering, valid validation methodology, meaningful metrics that address the hypothesis, clear interpretation of results and acknowledgment of limitations, and functional, error-free, and well-documented code that produces valid results. The prompt includes mandatory code checks requiring that code runs without errors, all imports and dependencies are properly handled, data loading and preprocessing are correct, statistical calculations are accurate, and output matches the described analysis. Additionally, the prompt enforces critical statistical terminology verification, ensuring that results are interpreted using proper language (e.g., “reject the null hypothesis” rather than “accept the null hypothesis”) and that p-values, effect sizes, and confidence intervals are properly reported. The prompt also includes a hypothesis alignment check, requiring that results not only show statistical significance but also that the direction of effect matches the stated hypothesis. The critic must output its verdict using XML (a markup language for storing and transporting data) tags: <verdict>NULL REJECTED</verdict>, <verdict>NULL NOT REJECTED</verdict>, or <verdict>NOT TESTABLE</verdict>, with the latter reserved for cases where dataset limitations discovered during analysis prevent meaningful hypothesis testing. If the critic agent is satisfied with the statistical results, it declares the hypothesis as verified. However, if it is not satisfied, the loop continues for a fixed number of inner loop iterations. After these inner iterations, the system initiates a new outer iteration, which resets the conversation context but incorporates summaries from previous attempts. The planning agent is then prompted to generate a new plan, enabling the system to explore alternative variables, statistical methods, or analytical systems. We note that even during the inner loop iterations, the system summarizes the conversation if it grows beyond a certain number of messages, to ensure the context window of the LLM is not exceeded.

The critic agent also includes a specialized debugging mode that activates automatically when the scientist agent’s code fails to execute. When code execution errors occur, the critic switches to a debugging query mode that focuses exclusively on identifying and resolving code issues rather than evaluating the hypothesis. In this mode, the critic analyzes execution failures, identifies likely causes (such as syntax errors, import issues, data access problems, statistical analysis errors, or data preprocessing issues), and provides specific, actionable feedback to guide the scientist toward a working solution. The debugging query emphasizes code robustness, error handling, and best practices, with particular attention to statistical analysis errors including array shape compatibility, missing value handling, sample size requirements, and data type conversions. Importantly, when in debugging mode, the critic does not provide a hypothesis verdict (NULL REJECTED/NULL NOT REJECTED/NOT TESTABLE) since the code has not successfully executed, allowing the conversation to continue after providing debugging guidance so the scientist can fix the code and reattempt the analysis.

### 2.4 Technical implementation and reproducibility

The workflow is orchestrated through a conversation management system that maintains the complete conversation history between agents, tracks turn counts and implements context window management. The conversation manager stores all messages in a structured format, including user inputs, agent responses, and system messages containing variable descriptions and dataset metadata. To prevent exceeding the LLM context window during extended conversations, the system implements automatic conversation truncation that preserves critical context: previous iteration summaries (for outer loop iterations), the initial hypothesis and preprocessing messages, variable description messages, and the most recent conversation exchanges. This selective preservation ensures that essential context is maintained while removing intermediate messages that may be redundant.

Dataset information is systematically provided to agents at multiple stages of the workflow to ensure continuous access to relevant context. During the preprocessing phase, the data preparation agent injects a comprehensive system message containing all available variables with their descriptions, data types, and dataset statistics (total rows, columns, missing values, numeric and categorical column counts). Additionally, before each scientist agent round within the inner loop, variable descriptions are automatically re-injected into the conversation context, providing fresh access to variable names, descriptions, and metadata. This repeated injection ensures that agents maintain awareness of available variables and their characteristics throughout extended conversations, even after conversation truncation. The truncation mechanism specifically preserves these variable description messages, recognizing their critical importance for informed analytical decision-making.

For outer loop iterations, conversation summaries are generated using the LLM to condense previous attempts into concise context. These summaries capture the hypothesis being tested, approaches attempted, variables used, results found, and reasons for non-approval, enabling subsequent iterations to avoid repeating failed approaches and explore alternative methodologies. The summary is prepended to the conversation context at the start of each new outer loop iteration, allowing the planning agent to adapt its strategy based on previous attempts.

The system includes a code executor with configurable timeout mechanisms (default: 30 seconds), persistent global namespaces for variable continuity across code blocks, and comprehensive error handling. All agents use the same LLM instance loaded in memory, each with its own specialized prompt. We primarily used the Qwen3-4B-Instruct model (Qwen/Qwen3-4B-Instruct-2507) as the client. The system tracks code execution outcomes and automatically switches the critic agent to debugging mode when code execution fails, allowing for targeted error resolution before hypothesis evaluation.

The system supports configuration management through YAML (a human-readable data serialization language) files with command-line overrides, enabling reproducible experiments with documented parameter settings. All runs include configuration logging, random seed management, and structured logging of experiment parameters for reproducibility. Configuration details including model name, seed, hypothesis mode, and other experiment parameters are automatically logged to JSON files for each run. All agent interactions are also logged to structured JSON files that capture the complete workflow: hypothesis details, preprocessing steps, conversation turns with agent responses, code execution results, statistical outputs, and final verdicts. This comprehensive logging enables full reproducibility and post-hoc analysis of the agentic decision-making process.

### 2.5 Data availability

Data from the NACC cohort can be requested and downloaded at https://naccdata.org.

## 3 RESULTS

We rigorously tested the multi-agentic AI system on the NACC cohort via an exemplar hypothesis evaluation, batch analyses of 100 curated ADRD hypotheses, scalability trials on subsampled datasets, and negative controls with non-ADRD queries. These assessments reveal the system’s proficiency in automated, verifiable hypothesis testing, with outcomes emphasizing its precision, adaptability, and domain-specific utility. Detailed results from each component are presented sequentially below.

In an exemplar application, the multi-agentic AI system processed the hypothesis “*Alcohol consumption is associated with increased dementia risk*” using the NACC cohort (**Figure 2**). The Data Preparation Agent selected 19 relevant variables, including alcohol measures (ALCFREQ, ALCOCCAS, ALCOHOL), demographics (NACCAGE, SEX, RACE, EDUC), dementia outcomes (NACCIDEM, NACCALZD, etc.), and comorbidities (DEP, HYPERT, STROKE); it replaced unknown values with NaN and expanded categorical values into a dataset with binary indicators for alcohol frequency. The Planning Agent specified chi-square tests with Cramer’s V to examine associations between alcohol frequency categories (less than monthly, monthly, weekly, few times per week, daily/almost daily) and binary dementia progression. Executing via listwise deletion, the Scientist Agent detected significant associations across all categories (p < 0.0001), with small effect sizes (Cramer’s V 0.040-0.049, highest at 0.049 for “few times per week”), implying a dose-response pattern. The Critic Agent affirmed methodological soundness, noting limitations like cross-sectional causality and recall bias, leading to null hypothesis rejection in a single iteration. This finding aligns with systematic reviews and cohort studies indicating that alcohol intake,^26–29^ even at moderate levels, elevates dementia risk without protective effects. Furthermore, analyses using NACC and similar data support heightened dementia risk from alcohol use disorder, particularly in subgroups like men with neuropsychiatric conditions.

**Figure 2:**
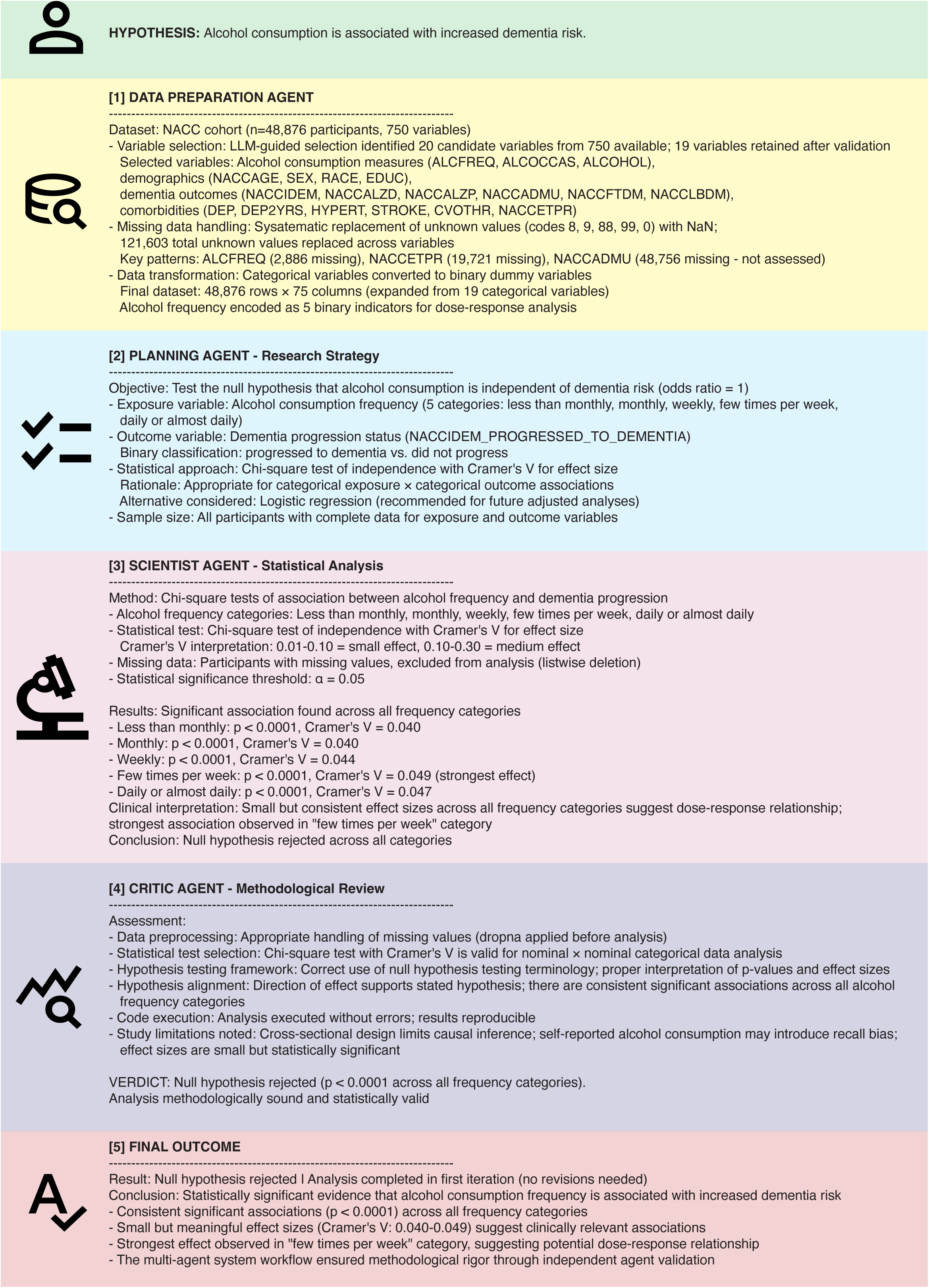
Sample execution log for a single hypothesis. This figure presents a detailed workflow log from the multi-agentic AI system evaluating the hypothesis “*Alcohol consumption is associated with increased dementia risk*” on the NACC cohort (n=48,876). It showcases sequential agent outputs: Data Preparation (variable selection, missing data handling, and transformation); Planning (research strategy with chi-square tests); Scientist (statistical execution revealing significant associations, p < 0.0001 across categories, Cramer’s V 0.040–0.049); Critic (methodological validation); and Final Outcome (null hypothesis rejection in one iteration), highlighting automated rigor in hypothesis verification.

We assessed the agentic system’s efficacy by applying it to 100 hypotheses curated from established ADRD literature (**Supplementary Table 1**), encompassing associations such as genetic risk factors (e.g., APOE ε4), lifestyle influences (e.g., physical activity), and biomarker interactions (e.g., amyloid and vascular pathology), using the NACC cohort (n=48,876 participants). As depicted in **Figure 3**, across replicated runs to account for variability in agentic processing, the system rejected the null hypothesis, indicating statistically significant verification of the proposed association, for a mean of 42.0±2.8 hypotheses, failed to reject the null (denoting absence of significant evidence) for 16.6±1.1 hypotheses, and classified 41.4±2.3 as not testable owing to missing variables, incompatible data formats, or insufficient cohort coverage. This outcome highlights the system’s capability to autonomously replicate a portion of known literature findings while prudently flagging unfeasible queries, thereby enhancing reliability in population-level analyses.

**Figure 3:**
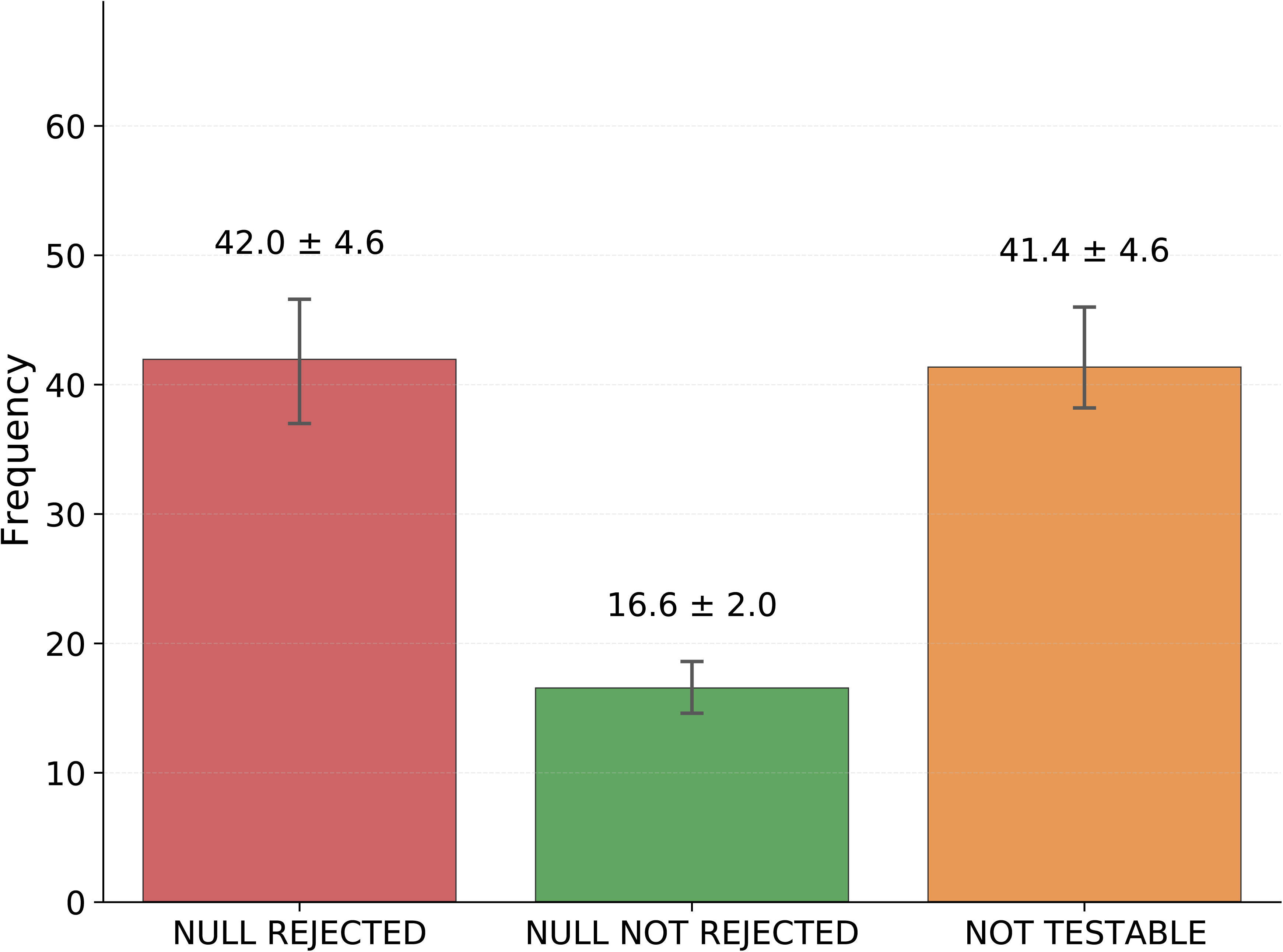
Performance of the agentic system on NACC data. Mean frequencies (± SD across multiple runs) for outcomes of 100 literature-derived ADRD hypotheses: null rejected (verified by significant association), null not rejected (no significant evidence), and not testable (due to data limitations).

To evaluate the scalability and sensitivity of the multi-agentic AI system to cohort size, we subsampled the NACC dataset (original n=48,876) to create reduced cohorts of 100, 1,000 and 10,000 participants while preserving demographic and clinical distributions, then applied the system to the same 100 literature-derived ADRD hypotheses as in prior analyses. As shown in **Figure 4**, performance exhibited a clear trend toward enhanced hypothesis verification with increasing dataset size: at n=100, the system rejected the null hypothesis for only 10.4 hypotheses (indicating limited detection of significant associations due to low power), failed to reject it for 31.6 (suggesting inconclusive evidence), and deemed 58.0 not testable owing to sparse or missing data in small samples. Scaling to n=1,000 improved resolution, with null rejections rising to 23.4, non-rejections at 27.4, and non-testable dropping to 49.2, reflecting better statistical power for identifying associations. At n=10,000, approaching the full cohort, the system achieved the highest verification rate, rejecting the null for 35.4 hypotheses, not rejecting for 21.6, and classifying 43.0 as not testable, demonstrating robustness in larger datasets where variable coverage and sample diversity enable more comprehensive analyses. Corresponding statistics between different groups are presented in **Supplementary Table 3**. These results underscore the system’s dependence on adequate data volume for accurate, reproducible hypothesis testing in ADRD research, with diminishing non-testability and stabilized outcomes as cohort size grows.

**Figure 4:**
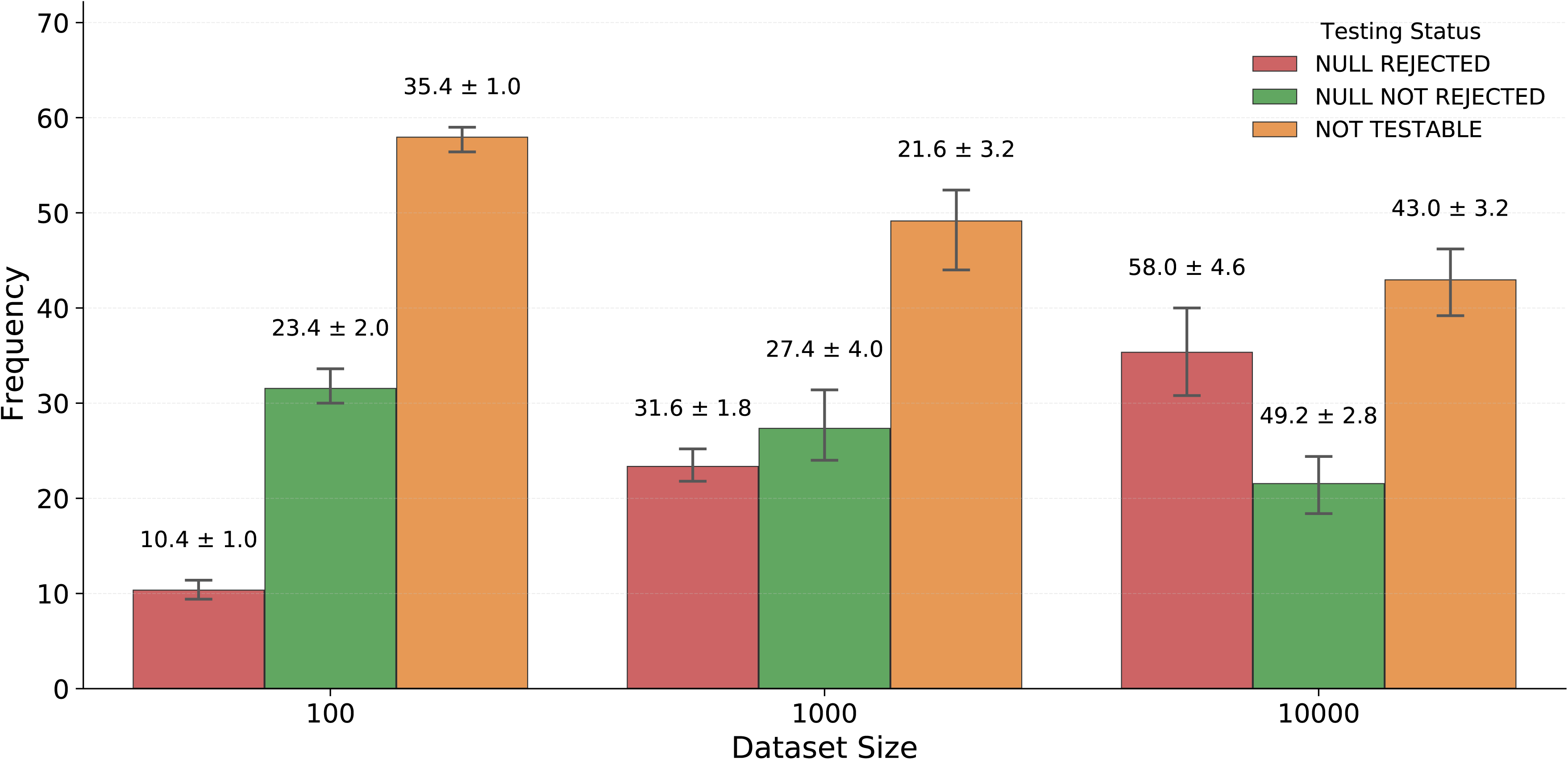
Scalability of the agentic system across dataset sizes. This plot demonstrates the mean frequencies of outcomes for 100 literature-derived ADRD hypotheses tested on subsampled NACC data at varying participant counts (100, 1,000, and 10,000 samples), categorized by null rejected (significant association verified), null not rejected (no significant evidence), and not testable (data limitations), highlighting improved verification rates with larger cohorts.

To assess the robustness of our agentic system in a control condition unrelated to ADRD, we evaluated its performance on 100 randomly generated hypotheses derived from general medical literature (**Supplementary Table 4**). These hypotheses spanned diverse topics such as cardiovascular disease, infectious diseases, endocrinology, and oncology, serving as a benchmark to test the system’s ability to handle queries outside its primary domain while providing a form of negative control validation. As illustrated in **Figure 5**, our system analyzed these hypotheses using NACC data. Across multiple runs, the system rejected the null hypothesis (indicating verification of the stated association) for an average of 7.6±0.8 hypotheses, failed to reject the null (indicating no significant evidence for the association) for 7.2±1.3 hypotheses, and deemed 85.2±0.6 hypotheses not testable due to insufficient or incompatible data in the NACC dataset. This high rate of non-testability validates the system’s conservative approach when applied to extraneous topics, highlighting its specificity for ADRD-related inquiries, minimizing spurious conclusions in unrelated contexts, and reducing the risk of false positives in out-of-domain scenarios. Although the hypotheses were randomly generated, few of them were still testable using the variables available in the NACC dataset. For example, the hypothesis “*Cholesterol-lowering diets reduce cardiovascular disease risk*” could be evaluated because the dataset included indicators of hypercholesterolemia, cardiac events (such as heart attack, angina, or bypass surgery), and medication use relevant to cholesterol and blood-pressure management. The presence of these well-defined clinical and behavioral variables enabled the construction of measurable exposure and outcome categories, making it possible to statistically assess at least some of the randomly produced hypotheses. Similarly, the hypothesis “*Obesity increases the risk of obstructive sleep apnea*” was also testable, as the dataset contained BMI category variables and a clearly defined indicator of diagnosed sleep apnea. These variables allowed for the creation of categorical exposure and outcome measures, making it feasible to examine the association statistically despite the hypothesis being randomly generated.

**Figure 5:**
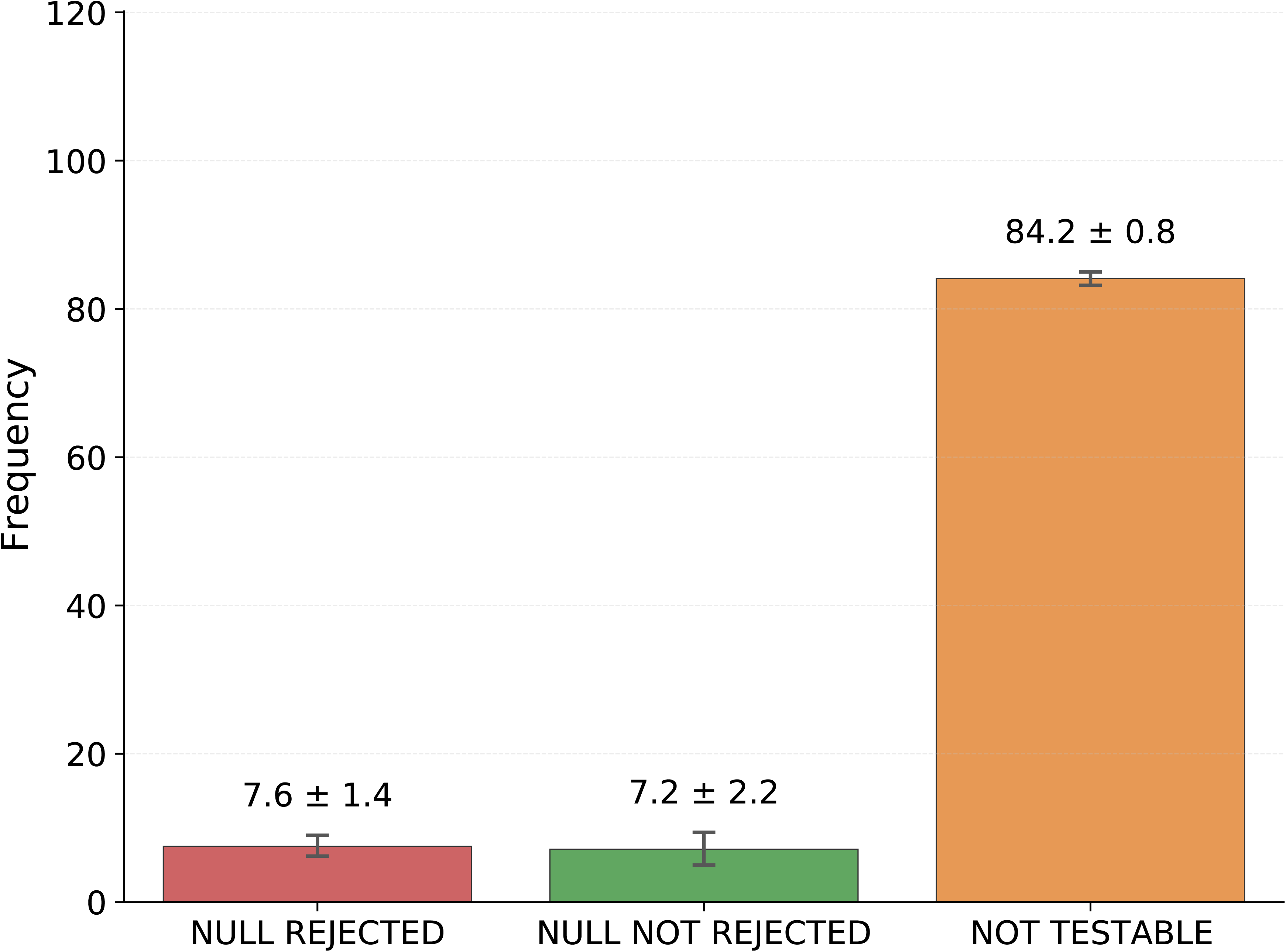
Performance of the agentic system in a negative control condition. This plot illustrates the performance of the agentic system on 100 randomly generated hypotheses from general medical literature (Supplementary Table 2) using NACC data. It shows the frequency of hypotheses where the null was rejected (indicating verification of the association), not rejected (indicating no significant evidence), and deemed not testable due to insufficient or incompatible data. The high non-testability rate serves as a negative control, validating the tool’s specificity and conservative approach for out-of-domain queries.

## 4 DISCUSSION

In this study, we developed and rigorously evaluated a multi-agentic AI system designed to automate and accelerate hypothesis testing in ADRD research. Using the NACC cohort (n=48,876 participants with rich demographic, clinical, neuropsychological, and biomarker data), we systematically tested 100 literature-derived ADRD hypotheses, their negated (opposite) counterparts, and critically, 100 randomly selected non-ADRD medical hypotheses as a negative control. The system’s specialized agents collaboratively handled the entire scientific workflow, i.e., from natural-language hypothesis interpretation, ontology-aware variable mapping, and missing-data handling, through statistical modeling, iterative critique, and reproducible result generation. Our results demonstrate high fidelity in replicating established ADRD associations while correctly identifying non-testable claims, maintaining logical consistency when evaluating negated hypotheses, showing robustness across dataset sizes from 100 to 10,000 participants, and exhibiting strong specificity by deeming most out-of-domain (non-ADRD) queries untestable.

Our multi-agentic system provides a structured approach to ADRD research by automating routine steps like data harmonization and basic testing, allowing researchers to focus more on interpretation and follow-up. It could help connect teams with varying resources, encouraging broader participation in examining disease pathways. For instance, midlife hypertension’s adverse impact on cognitive decline or stroke history’s link to dementia. By integrating with emerging platforms such as the AD Workbench along with Findable, Accessible, Interoperable, and Reusable (FAIR) principles, it might facilitate blending datasets for cross-study comparisons, such as metabolic syndrome’s role in cognitive decline or sleep disorders’ ties to amyloid buildup, fostering collaborative efforts to address dementia’s multifactorial aspects. Over time, this could contribute to practical applications in biomarker discovery or treatment evaluation, making analyses more inclusive across settings.

Our study has a few limitations. The hypotheses were selected from existing literature, so outcomes may align closely with those sources rather than highlight novel patterns, and the NACC dataset’s emphasis could underrepresent certain groups, such as specific ethnicities or regions. Dependence on large language models (e.g., Qwen3-4B-Instruct-2507) might carry over training biases, influencing interpretations in some areas. The iterative loops addressed many scenarios but could overlook nuances in complex cases, such as mixed dementia etiologies or visuospatial issues in Lewy body dementia. Broader challenges include data diversity, where training biases might constrain applicability, and ethical considerations around AI in health research call for ongoing human oversight and real-world validation.

In conclusion, these findings establish the agentic system as a reliable, transparent, and scalable tool that substantially reduces manual effort and analytic variability in population-level dementia research. This system also provides a basis for integrating AI into ADRD studies, connecting datasets with systematic explorations. Future developments could incorporate federated learning for multi-site privacy or adaptable agents for new biomarkers from wearables. Through such steps, these tools might support progress in understanding dementia’s diverse components, from genetic and lifestyle elements to therapeutic responses, aiding shared initiatives for improved outcomes.

## Data Availability

All data produced in the present study are available upon reasonable request to the authors.

## ACKNOWLEDGMENTS

The manuscript was originally drafted without the use of AI technologies. Subsequently, our agentic system was used to edit some portions of the manuscript. Finally, the authors reviewed and edited the content as needed and take full responsibility for the content of the publication.

The NACC database is funded by NIA/NIH Grant U24 AG072122. NACC data are contributed by the NIA-funded ADRCs: P30 AG062429 (PI James Brewer, MD, PhD), P30 AG066468 (PI Oscar Lopez, MD), P30 AG062421 (PI Bradley Hyman, MD, PhD), P30 AG066509 (PI Thomas Grabowski, MD), P30 AG066514 (PI Mary Sano, PhD), P30 AG066530 (PI Helena Chui, MD), P30 AG066507 (PI Marilyn Albert, PhD), P30 AG066444 (PI David Holtzman, MD), P30 AG066518 (PI Lisa Silbert, MD, MCR), P30 AG066512 (PI Thomas Wisniewski, MD), P30 AG066462 (PI Scott Small, MD), P30 AG072979 (PI David Wolk, MD), P30 AG072972 (PI Charles DeCarli, MD), P30 AG072976 (PI Andrew Saykin, PsyD), P30 AG072975 (PI Julie A. Schneider, MD, MS), P30 AG072978 (PI Ann McKee, MD), P30 AG072977 (PI Robert Vassar, PhD), P30 AG066519 (PI Frank LaFerla, PhD), P30 AG062677 (PI Ronald Petersen, MD, PhD), P30 AG079280 (PI Jessica Langbaum, PhD), P30 AG062422 (PI Gil Rabinovici, MD), P30 AG066511 (PI Allan Levey, MD, PhD), P30 AG072946 (PI Linda Van Eldik, PhD), P30 AG062715 (PI Sanjay Asthana, MD, FRCP), P30 AG072973 (PI Russell Swerdlow, MD), P30 AG066506 (PI Glenn Smith, PhD, ABPP), P30 AG066508 (PI Stephen Strittmatter, MD, PhD), P30 AG066515 (PI Victor Henderson, MD, MS), P30 AG072947 (PI Suzanne Craft, PhD), P30 AG072931 (PI Henry Paulson, MD, PhD), P30 AG066546 (PI Sudha Seshadri, MD), P30 AG086401 (PI Erik Roberson, MD, PhD), P30 AG086404 (PI Gary Rosenberg, MD), P20 AG068082 (PI Angela Jefferson, PhD), P30 AG072958 (PI Heather Whitson, MD), P30 AG072959 (PI James Leverenz, MD).

## DECLARATIONS

V.B.K. is a co-founder and equity holder of deepPath Inc. and Cognimark, Inc. He also serves on the scientific advisory board of Altoida Inc. The remaining authors declare no competing interests.

