## Supplementary Table 1 for "Agentic AI for automated hypothesis testing in Alzheimer’s disease and related dementias"

**Supplementary Table 1: Literature-derived hypotheses.** This table lists 100 hypotheses derived from literature along with links to corresponding citations on topics pertaining to Alzheimer’s disease and related dementias.

| **No.** | | **Hypothesis** | **Citation** | | **Link** |
| --- | --- | --- | --- | --- | --- |
| **1** | | Higher age is associated with increased dementia incidence. | The incidence of dementia rises exponentially to the age of 90 years. | | <https://pubmed.ncbi.nlm.nih.gov/9748017/> |
|  |  |  | In the pooled analysis, the incidence rates increased with age without any substantial difference between men and women. | | <https://pubmed.ncbi.nlm.nih.gov/10854355/> |
|  |  |  | The overall incidence rate of all-cause dementia was 18.2% (95% confidence interval [CI], 15.3-21.5) per year and was similar for men and women (risk ratio, 0.94; 95% CI, 0.65–1.37). Rates increased exponentially with age from 12.7% per year in the 90–94-year age group, to 21.2% per year in the 95–99-year age group, to 40.7% per year in the 100+-year age group. | | <https://pmc.ncbi.nlm.nih.gov/articles/PMC3385995/> |
| **2** | | Greater years of education are linked to lower dementia incidence (cognitive reserve). | In this study an inverse dose-response relation was found between education and dementia--in particular, Alzheimer's disease. | | <https://pubmed.ncbi.nlm.nih.gov/7728032/> |
|  |  |  | These results support the theory that individuals with greater cognitive reserve, as reflected in years of education, are better able to cope with AD brain pathology without observable deficits in cognition. | | <https://pubmed.ncbi.nlm.nih.gov/17224578/> |
|  |  |  | Participants without formal education presented more than double the prevalence of dementia (21.37%) compared to those with at least one year of formal education (9.88%). Studies with more recent data collection showed higher dementia prevalence. | | <https://pubmed.ncbi.nlm.nih.gov/35931410/> |
| **3** | | Women are at higher risk of developing dementia than men. | The odds ratios for women to develop incidence of dementia and AD relative to men are 1.18 (95% confidence interval, 0.95-1.46) and 1.56 (95% confidence interval, 1.16-2.10), respectively. | | <https://pubmed.ncbi.nlm.nih.gov/9736007/> |
|  |  |  | The pooled risk of all-cause dementia was higher in women than men after adjusting for age and education (HR, 1.12 [1.02, 1.23]), although not uniform across the individual cohorts. | | <https://pubmed.ncbi.nlm.nih.gov/36790027/> |
|  |  |  | These results suggest that women have greater cognitive reserve but faster later-life cognitive decline than men. Evidence suggests that dementia incidence in Europe and the US has declined over the past 25 years, but declines were less in women than in men. Our findings suggest that women are at risk for delayed identification of cognitive decline, yet more rapid trajectory of decline, suggesting increased risk of dementia and disability compared with men, consistent with research showing that women with mild cognitive impairment or AD have faster cognitive decline than men. Women may thus have greater needs for caregiving and functional support resources, particularly given women’s longer life expectancy compared with men. | | <https://pubmed.ncbi.nlm.nih.gov/33630089/> |
| **4** | | APOE ε4 carriage increases risk for cognitive impairment. | The APOE epsilon4 allele represents a major risk factor for AD in all ethnic groups studied, across all ages between 40 and 90 years, and in both men and women. | | <https://pubmed.ncbi.nlm.nih.gov/9343467/> |
|  |  |  | The apolipoprotein E type 4 allele (APOE-epsilon 4) is genetically associated with the common late onset familial and sporadic forms of Alzheimer's disease (AD). Risk for AD increased from 20% to 90% and mean age at onset decreased from 84 to 68 years with increasing number of APOE-epsilon 4 alleles in 42 families with late onset AD. Thus APOE-epsilon 4 gene dose is a major risk factor for late onset AD and, in these families, homozygosity for APOE-epsilon 4 was virtually sufficient to cause AD by age 80. | | <https://pubmed.ncbi.nlm.nih.gov/8346443/> |
|  |  |  | The APOE-ε4 allele is associated with a moderately increased risk for progression from MCI to AD-type dementia. | | <https://pubmed.ncbi.nlm.nih.gov/21493755/> |
| **5** | | Hispanic ethnicity shows attenuated APOE ε4 effect compared with non-Hispanics. | Middle-aged Hispanic APOE ε4 carriers have higher in vivo brain amyloid burden compared with noncarriers, as reported in non-Hispanics. | | <https://pubmed.ncbi.nlm.nih.gov/32847955/> |
|  |  |  | Apolipoprotein E epsilon4 had a consistent lowering effect on age at onset of FAD, but this was attenuated in SAD. | | <https://pubmed.ncbi.nlm.nih.gov/17101827/> |
|  |  |  | The APOE epsilon4-AD association was weaker among African Americans and Hispanics, but there was significant heterogeneity in ORs among studies of African Americans (P<.03). | | <https://pubmed.ncbi.nlm.nih.gov/9343467/> |
| **6** | | Married status is associated with lower risk of dementia vs. unmarried/widowed. | Being married is associated with reduced risk of dementia than widowed and lifelong single people, who are also underdiagnosed in routine clinical practice. | | <https://pubmed.ncbi.nlm.nih.gov/29183957/> |
|  |  |  | People cohabiting with a partner in mid-life (mean age 50.4) were less likely than all other categories (single, separated, or widowed) to show cognitive impairment later in life at ages 65-79. | | <https://pubmed.ncbi.nlm.nih.gov/19574312/> |
|  |  |  | The relative risks (RRs) of dementia (RR = 1.91, p = 0.018) and of AD (RR = 2.68, p<0.001) were increased for the never-married individuals compared with those who were married or cohabitants. | | <https://pubmed.ncbi.nlm.nih.gov/10599764/> |
| **7** | | MoCA is more sensitive than MMSE for detecting MCI. | MoCA test better meets the criteria for screening tests for the detection of MCI among patients over 60 years of age than MMSE. | | <https://pubmed.ncbi.nlm.nih.gov/27992895/> |
|  |  |  | The MoCA had better performance than the other MCI screening tests. | | <https://pubmed.ncbi.nlm.nih.gov/26052687/> |
|  |  |  | Using a cutoff score 26, the MMSE had a sensitivity of 18% to detect MCI, whereas the MoCA detected 90% of MCI subjects. In the mild AD group, the MMSE had a sensitivity of 78%, whereas the MoCA detected 100%. | | <https://pubmed.ncbi.nlm.nih.gov/15817019/> |
| **8** | | Higher CDR-Sum of Boxes scores are associated with greater dementia severity. | The CDR-SOB score compares well with the global CDR score for dementia staging. Owing to the increased range of values, the CDR-SOB score offers several advantages over the global score, including increased utility in tracking changes within and between stages of dementia severity. Interpretive guidelines for CDR-SOB scores are provided. | | <https://pubmed.ncbi.nlm.nih.gov/18695059/> |
|  |  |  | The previously proposed CDR-SB ranges successfully classified the vast majority of patients across all impairment ranges with a kappa of 0.91 and 94% overall correct classification rate. | | <https://pubmed.ncbi.nlm.nih.gov/20558394/> |
|  |  |  | A longitudinal increase (P < .0001) in CDR-SB was observed. The annual rate of change in CDR-SB scores was 1.43 (standard error [SE] = 0.05) in the CDR 0.5 sample and 1.91 (SE = 0.07) in the CDR 1 sample. | | <https://pubmed.ncbi.nlm.nih.gov/22858530/> |
| **9** | | Diabetes is associated with increased risk of cognitive impairment. | During the follow-up, 126 patients became demented, of whom 89 had AD. Diabetes mellitus almost doubled the risk of dementia | | <https://pubmed.ncbi.nlm.nih.gov/10599761/> |
|  |  |  | These findings provide evidence that the association between type 2 diabetes and dementia/CIND among Mexican Americans remains strong after accounting for competing risk of mortality. | | <https://pubmed.ncbi.nlm.nih.gov/23514732/> |
|  |  |  | Diabetes characterized by poor glycemic control or cardiovascular complications is related to a greater risk of the development and progression of cognitive impairment. | | <https://pubmed.ncbi.nlm.nih.gov/34636485/> |
| **10** | | Hypertension in midlife increases risk of cognitive decline. | In this community-based cohort with long-term follow-up, sustained hypertension in midlife to late life and a pattern of midlife hypertension and late-life hypotension, compared with midlife and late-life normal BP, were associated with increased risk for subsequent dementia. | | <https://pubmed.ncbi.nlm.nih.gov/31408138/> |
|  | |  | Elevated blood pressure during midlife, persistence of elevated blood pressure into late life, and, among nonhypertensives, a steep decline in blood pressure during mid- to late life were associated with an increased dementia risk in a community-based cohort. | | <https://pubmed.ncbi.nlm.nih.gov/29117954/> |
|  | |  | Findings suggest that cumulative BP over the course of midlife predicts risk of dementia in later life. | | <https://pubmed.ncbi.nlm.nih.gov/37394941/> |
| **11** | | Prior stroke increases risk of dementia. | After study methods and case mix are taken into account, reported estimates of the prevalence of dementia are consistent: 10% of patients had dementia before first stroke, 10% developed new dementia soon after first stroke, and more than a third had dementia after recurrent stroke. The strong association of post-stroke dementia with multiple strokes and the prognostic value of other stroke characteristics highlight the central causal role of stroke itself as opposed to the underlying vascular risk factors and, thus, the likely effect of optimum acute stroke care and secondary prevention in reducing the burden of dementia. | | <https://pubmed.ncbi.nlm.nih.gov/19782001/> |
|  |  |  | Risk of dementia increased with both the number and severity of strokes. Compared with no stroke, risk of dementia by adjusted hazard ratio was 1.76 (95% CI, 1.49-2.00) for 1 minor to mild stroke, 3.47 (95% CI, 2.23-5.40) for 1 moderate to severe stroke, 3.48 (95% CI, 2.54-4.76) for 2 or more minor to mild strokes, and 6.68 (95% CI, 3.77-11.83) for 2 or more moderate to severe strokes. | | <https://pubmed.ncbi.nlm.nih.gov/35072712/> |
|  |  |  | his study confirms the high prevalence of PSCI in diverse populations, highlights common risk factors, in particular diabetes mellitus, and points to ethnoracial differences that warrant attention in the development of prevention strategies. | | <https://pubmed.ncbi.nlm.nih.gov/31712368/> |
| **12** | | Depression is associated with an increased risk of developing dementia. | The hazard of dementia among those diagnosed with depression was 2.41 times that of the comparison cohort (95% CI, 2.35-2.47).... Results suggest that the risk of dementia was more than doubled for both men and women with diagnosed depression. The persistent association between dementia and depression diagnosed in early and middle life suggests that depression may increase dementia risk. | | <https://pubmed.ncbi.nlm.nih.gov/37486689/> |
|  |  |  | The adjusted hazard of dementia was increased by approximately 20% for midlife depressive symptoms only (hazard ratio, 1.19 [95% CI, 1.07-1.32]), 70% for late-life symptoms only (1.72 [1.54-1.92]), and 80% for both (1.77 [1.52-2.06])....Depressive symptoms in midlife or in late life are associated with an increased risk of developing dementia. | | <https://pubmed.ncbi.nlm.nih.gov/22566581/> |
|  |  |  | Depression was associated with a 51% higher risk of dementia, among which the increasing, chronically high, and chronically low courses were associated with increased dementia risk, while no association was found in the decreasing course. | | <https://pubmed.ncbi.nlm.nih.gov/36526487/> |
| **13** | | Anxiety disorders are linked to cognitive decline. | A bidirectional relationship with a 2-year lag between anxiety and MCI was mediated through perceived stress. | | <https://pubmed.ncbi.nlm.nih.gov/36855309/> |
|  |  |  | Depression, anxiety, and apathy symptoms are highly prevalent across dementia stages. | | <https://pubmed.ncbi.nlm.nih.gov/33905138/> |
|  |  |  | These results support a unique pathophysiological relationship between anxiety and AD that can be reflected in plasma biomarkers, suggestive of heightened neurodegeneration. | | <https://pubmed.ncbi.nlm.nih.gov/39604275/> |
| **14** | | Smoking increases risk of Alzheimer’s disease. | The available data indicate that smoking is a significant risk factor for AD. | | <https://pubmed.ncbi.nlm.nih.gov/20110594/> |
|  |  |  | Smoking was associated with a doubling of the risk of dementia and Alzheimer's disease. | | <https://pubmed.ncbi.nlm.nih.gov/9652667/> |
|  |  |  | In this large cohort, heavy smoking in midlife was associated with a greater than 100% increase in risk of dementia, AD, and VaD more than 2 decades later. | | <https://pubmed.ncbi.nlm.nih.gov/20975015/> |
| **15** | | Alcohol consumption is associated with increased dementia risk. | This study identified a positive linear causal relationship between alcohol consumption and dementia among current drinkers. | | <https://pubmed.ncbi.nlm.nih.gov/39290634/> |
|  |  |  | Alcohol use disorders were a major risk factor for onset of all types of dementia, and especially early-onset dementia. | | <https://pubmed.ncbi.nlm.nih.gov/29475810/> |
|  |  |  | The findings of this study suggest that alcohol-induced loss of consciousness, irrespective of overall alcohol consumption, is associated with a subsequent increase in the risk of dementia. | | <https://pubmed.ncbi.nlm.nih.gov/32902651/> |
| **16** | | Hypercholesterolemia in midlife is associated with higher risk of dementia. | Midlife serum total cholesterol was associated with an increased risk of AD and VaD. Even moderately elevated cholesterol increased dementia risk. | | <https://pubmed.ncbi.nlm.nih.gov/19648749/> |
|  |  |  | Relative risk of developing AD for adults with high TC in midlife was 2.14 (95% CI 1.33-3.44) compared with normal cholesterol. | | <https://pubmed.ncbi.nlm.nih.gov/27911314/> |
|  |  |  | Midlife hypercholesterolemia was associated with increased incidence of mild cognitive impairment (effect size [ES] = 2.01; 95% confidence interval [CI] 1.19 to 2.84; I2 = 0.0%) and all-cause dementia (ES = 1.14; 95% CI: 1.07 to 1.21; I2 = 0.0%). | | <https://pubmed.ncbi.nlm.nih.gov/36911359/> |
| **17** | | Myocardial infarction is associated with cognitive decline. | This cohort study using pooled data from 6 cohort studies found that incident MI was not associated with a decrease in global cognition, memory, or executive function at the time of the event compared with no MI but was associated with faster declines in global cognition, memory, and executive function over time. | | <https://pubmed.ncbi.nlm.nih.gov/37252710/> |
|  |  |  | MI was associated with higher risk of vascular dementia throughout follow-up, and this association was stronger in patients with stroke. | | <https://pubmed.ncbi.nlm.nih.gov/29025764/> |
|  |  |  | Incident CHD is associated with accelerated cognitive decline after, but not before, the event. | | <https://pubmed.ncbi.nlm.nih.gov/31221251/> |
| **18** | | Lewy body dementia shows symptoms of visual hallucinations. | Impairment of visual-spatial and perceptual abilities in DLB represents a disease related cognitive signature, independent of the presence of VHs, for which it may represent a predisposing condition. | | <https://pubmed.ncbi.nlm.nih.gov/23264688/> |
|  |  |  | Thus, our results suggest that the distribution of temporal lobe LB is more related to the presence and duration of visual hallucinations in cases with LB than to the presence, severity or duration of dementia. | | <https://pubmed.ncbi.nlm.nih.gov/11844739/> |
|  |  |  | These findings support the hodotopic hypothesis of VH and may reflect a link between VH phenomenology, LBD neuropathological progression and the involvement of specific neurotransmitter systems. | | <https://pubmed.ncbi.nlm.nih.gov/35099586/> |
| **19** | | Mixed dementia is common in older adults. | In persons with dementia, over 50% had multiple diagnoses (AD, PD/LBD, or infarcts), whereas, in persons without dementia, over 80% had one or no diagnosis. After accounting for age, persons with multiple diagnoses were almost three times (OR = 2.8; 95% CI = 1.2, 6.7) more likely to exhibit dementia compared to those with one pathologic diagnosis. | | <https://pubmed.ncbi.nlm.nih.gov/17568013/> |
|  |  |  | Neuropathology was ubiquitous, with 94% of participants having 1+, 78% having 2+, 58% having 3+, and 35% having 4+. AD was most frequent (65%) but rarely occurred in isolation (9%). | | <https://pubmed.ncbi.nlm.nih.gov/29244218/> |
|  |  |  | Older persons with AD pathology, often have concomitant cerebrovascular disease pathologies (macroinfarcts, microinfarcts, atherosclerosis, arteriolosclerosis, cerebral amyloid angiopathy) as well as other concomitant neurodegenerative disease pathologies (Lewy bodies, TDP-43, hippocampal sclerosis). | | <https://pubmed.ncbi.nlm.nih.gov/28488154/> |
| **20** | | APOE ε4 carriers show earlier amyloid accumulation. | Together, our results demonstrate that apoE4 has the greatest impact on amyloid during the seeding stage, likely by perturbing Aβ clearance and enhancing Aβ aggregation. | | <https://pubmed.ncbi.nlm.nih.gov/29216449/> |
|  |  |  | The findings in this study indicate that amyloid-related tau accumulation was accelerated in ApoE4 carriers at lower amyloid levels, suggesting that ApoE4 may facilitate earlier amyloid-driven tau spreading across connected brain regions. | | <https://pubmed.ncbi.nlm.nih.gov/37930695/> |
|  |  |  | The age at which 15% of the participants with normal cognition were amyloid positive was approximately 40 years for APOE ε4ε4 carriers, 50 years for ε2ε4 carriers, 55 years for ε3ε4 carriers, 65 years for ε3ε3 carriers, and 95 years for ε2ε3 carriers. | | <https://pubmed.ncbi.nlm.nih.gov/25988462/> |
| **21** | | Obesity in midlife increases risk of late-life dementia. | Both overweight and obesity at midlife independently increase the risk of dementia, AD, and VaD. | | <https://pubmed.ncbi.nlm.nih.gov/21536637/> |
|  |  |  | In evaluations of midlife obesity, an increased risk of dementia was found for obese (BMI >30) vs normal-weight (BMI 20-25) persons, adjusted for demographics (hazard ratio [HR], 1.39; 95% confidence interval [CI], 1.03-1.87) and for cardiovascular risk factors (1.36; 0.94-1.95). | | <https://pubmed.ncbi.nlm.nih.gov/19273752/> |
|  |  |  | Obesity in middle age increases the risk of future dementia independently of comorbid conditions. | | <https://pubmed.ncbi.nlm.nih.gov/15863436/> |
| **22** | | Metabolic syndrome is associated with cognitive decline. | The MetS was associated with an increased incidence of MCI and progression to dementia. | | <https://pubmed.ncbi.nlm.nih.gov/26926205/> |
|  |  |  | MetS at baseline was associated with an increased risk of cognitive decline on MMSE (hazard ratio [HR] = 1.22 [1.08-1.37]; p = 0.001) and BVRT (HR = 1.13 [1.01-1.26]; p = 0.03) but not on IST (HR = 1.11 [0.95-1.29]; p = 0.18). Among MetS components, hypertriglyceridemia and low HDL cholesterol were significantly associated with higher decline on MMSE; diabetes, but not elevated fasting glycemia, was significantly associated with higher decline on BVRT and IST. | | <https://pubmed.ncbi.nlm.nih.gov/21288982/> |
|  |  |  | These findings support the hypothesis that the metabolic syndrome contributes to cognitive impairment in elders, but primarily in those with high level of inflammation. | | <https://pubmed.ncbi.nlm.nih.gov/15536110/> |
| **23** | | Systemic inflammation is associated with cognitive decline | After adjusting for demographic variables, vascular risk factors, and comorbidities, each standard deviation (SD) increase in midlife inflammation composite score was associated with an additional 20-year decline of -0.035 SD (95% confidence interval: -0.062 to -0.007) on the cognitive composite score. | | <https://pubmed.ncbi.nlm.nih.gov/30760633/> |
|  |  |  | With data collected over 20 years, this study demonstrated greater likelihood of cognitive impairment in individuals with repeated high or increasing IL-6. | | <https://pubmed.ncbi.nlm.nih.gov/25123210/> |
|  |  |  | Our results suggest that low-grade inflammation in midlife is an independent risk factor for poorer cognitive performance later in life. | | <https://pubmed.ncbi.nlm.nih.gov/36195448/> |
| **24** | | Sleep disturbances are linked to cognitive impairment. | Persistent short sleep duration at age 50, 60, and 70 compared to persistent normal sleep duration was also associated with a 30% increased dementia risk independently of sociodemographic, behavioural, cardiometabolic, and mental health factors. These findings suggest that short sleep duration in midlife is associated with an increased risk of late-onset dementia. | | <https://pubmed.ncbi.nlm.nih.gov/33879784/> |
|  |  |  | Among 1,443 subjects, 69(4.78%) had MCI, and 830 (57.52%) had sleep disturbance. In bivariate analysis, MCI was associated with sleep disturbance (ρ = 0.094, P<0.001). | | <https://pubmed.ncbi.nlm.nih.gov/36320021/> |
|  |  |  | Ten types of sleep conditions or parameters, including six (insomnia, fragmentation, daytime dysfunction, prolonged latency, rapid eye movement sleep behaviour disorder and excessive time in bed) with moderate-to-high levels of evidence, were linked to higher risk of all-cause cognitive disorders. Furthermore, a U-shaped relationship was revealed for the associations with sleep duration. | | <https://pubmed.ncbi.nlm.nih.gov/31879285/> |
| **25** | | Greater physical activity lowers risk of cognitive decline. | Meta-analysis, using the quality-effects model, suggests that participants with higher levels of physical activity, when compared to those with lower levels, are at reduced risk of cognitive decline, RR 0.65, 95% CI 0.55-0.76, and dementia, RR 0.86, 95% CI 0.76-0.97. | | <https://pubmed.ncbi.nlm.nih.gov/24885250/> |
|  |  |  | The cumulative analysis for all the studies under a random-effects model showed that subjects who performed a high level of physical activity were significantly protected (-38%) against cognitive decline during the follow-up (hazard ratio (HR) 0.62, 95% confidence interval (CI) 0.54-0.70; P < 0.00001). Furthermore, even analysis of low-to-moderate level exercise also showed a significant protection (-35%) against cognitive impairment (HR 0.65, 95% CI 0.57-0.75; P < 0.00001). | | <https://pubmed.ncbi.nlm.nih.gov/20831630/> |
|  |  |  | PA was associated with a decreased risk of all-cause dementia (pooled relative risk 0.80, 95% CI 0.77 to 0.84, n=257 983), Alzheimer's disease (0.86, 95% CI 0.80 to 0.93, n=128 261) and vascular dementia (0.79, 95% CI 0.66 to 0.95, n=33 870), even in longer follow-ups (≥20 years) for all-cause dementia and Alzheimer's disease. | | <https://pubmed.ncbi.nlm.nih.gov/35301183/> |
| **26** | | Mediterranean diet adherence reduces risk of MCI and progression to dementia. | Higher adherence to MeD was associated with a lower likelihood of ICI independent of potential confounders. | | <https://pubmed.ncbi.nlm.nih.gov/23628929/> |
|  |  |  | Higher adherence to the MeDi is associated with a trend for reduced risk of developing MCI and with reduced risk of MCI conversion to AD. | | <https://pubmed.ncbi.nlm.nih.gov/19204158/> |
|  |  |  | In an older population, a Mediterranean diet supplemented with olive oil or nuts is associated with improved cognitive function. | | <https://pubmed.ncbi.nlm.nih.gov/25961184/> |
| **27** | | Low CSF Aβ42 and high tau identify AD pathology and predict progression. | Aβ42 was the strongest predictor of progression to MCI or AD with an adjusted hazard ratio (HR) of 16.0 (3.8-66.4). | | <https://pubmed.ncbi.nlm.nih.gov/23232269/> |
|  |  |  | We found increased CSF-tau and decreased CSF-Abeta42 levels in probable and possible AD. Sensitivity was 94% for probable AD, 88% for possible AD, and 75% for mild cognitive impairment, whereas specificity was 100% for psychiatric disorders and 89% for nondemented. | | <https://pubmed.ncbi.nlm.nih.gov/11255440/> |
|  |  |  | Using age-matched controls with normal CSF T/Abeta we showed that the high CSF T/Abeta subgroup of controls had significantly increased frequency of the epsilon4 allele of the apolipoprotein E gene and significantly increased risk of conversion to MCI during follow-up of up to 42 months suggesting that they had latent AD at the time of lumbar puncture. | | <https://pubmed.ncbi.nlm.nih.gov/17698783/> |
| **28** | | Amyloid PET positivity predicts faster cognitive decline and conversion to AD. | PET regional binding patterns are consistent with known neuropathologic patterns of plaque and tangle brain accumulation, spreading from the medial temporal to other neocortical regions as disease progresses. Because binding patterns predict future cognitive decline and increase over time along with clinical decline, [(18)F]FDDNP PET scanning may have practical utility in identifying people at risk for future cognitive decline and in tracking the effectiveness of novel interventions designed to prevent or delay neurodegeneration and cognitive decline. | | <https://pubmed.ncbi.nlm.nih.gov/22332188/> |
|  |  |  | Overall, these findings suggest that in CN, MCI and AD subjects, florbetapir PET Aβ+ subjects show greater cognitive and global deterioration over a 3-year follow-up than Aβ- subjects do. | | <https://pubmed.ncbi.nlm.nih.gov/24614494/> |
|  |  |  | PIB-positive subjects with mild cognitive impairment (MCI) are significantly more likely to convert to AD than PIB-negative patients, faster converters having higher PIB retention levels at baseline than slower converters. | | <https://pubmed.ncbi.nlm.nih.gov/19587325/> |
| **29** | | Hippocampal atrophy on MRI predicts conversion from MCI to AD. | Smaller hippocampi and specifically CA1 and subicular involvement are associated with increased risk for conversion from MCI to AD. | | <https://pubmed.ncbi.nlm.nih.gov/16682538/> |
|  |  |  | In older patients with MCI, hippocampal atrophy determined by premorbid MRI-based volume measurements is predictive of subsequent conversion to AD. | | <https://pubmed.ncbi.nlm.nih.gov/10227624/> |
|  |  |  | Smaller hippocampal and entorhinal cortex volumes each contribute to the prediction of conversion to Alzheimer disease. | | <https://pubmed.ncbi.nlm.nih.gov/17353470/> |
| **30** | | AD cortical thinning pattern predicts AD. | Thinning in specific cortical areas known to be affected by Alzheimer disease (AD) is detectable in individuals with questionable AD dementia (QAD) and predicts conversion to mild AD dementia. This method could be useful for identifying individuals at relatively high risk for imminent progression from QAD to mild AD dementia, which may be of value in clinical trials. | | <https://pubmed.ncbi.nlm.nih.gov/19109536/> |
|  |  |  | In presymptomatic individuals harboring indicators of AD, baseline thickness in AD-vulnerable cortical regions was significantly reduced compared with that of healthy control individuals, but baseline hippocampal volume was not. | | <https://pubmed.ncbi.nlm.nih.gov/21825241/> |
|  |  |  | Cortical thickness patterns can reflect pathophysiological and clinical changes in AD. | | <https://pubmed.ncbi.nlm.nih.gov/27239533/> |
| **31** | | White matter hyperintensities are associated with higher risk of dementia. | White matter hyperintensities portended an increased risk of stroke, amnestic mild cognitive impairment, dementia, and death independent of vascular risk factors and interim vascular events. | | <https://pubmed.ncbi.nlm.nih.gov/20167919/> |
|  |  |  | White matter lesions, especially in the periventricular region, increase the risk of dementia in elderly people. | | <https://pubmed.ncbi.nlm.nih.gov/15477506/> |
|  |  |  | A greater severity of periventricular white-matter lesions was also associated with an increased risk of dementia....Furthermore, we showed that a greater severity of periventricular white-matter lesions, also thought to result from small-vessel disease, was associated with an increased risk of dementia. | | <https://pubmed.ncbi.nlm.nih.gov/12660385/> |
| **32** | | Greater white matter hyperintensities burden predicts faster cognitive decline in PD as well. | In this cohort study, higher levels of both WMH and indices of cerebrovascular disease pathologies in aging brains were associated with more rapid progressive parkinsonism. | | <https://pubmed.ncbi.nlm.nih.gov/34724033/> |
|  |  |  | This study demonstrated that the burden of WMH within cholinergic pathways was significantly higher in patients with PDD relative to other groups, and that cholinergic WMH was significantly correlated with a decline in frontal executive function and attention. | | <https://pubmed.ncbi.nlm.nih.gov/22228726/> |
|  |  |  | The frequency and the extent of periventricular hyperintensities were significantly higher in patients with PD than in healthy subjects. Moreover, within the PD group, the patients who had periventricular hyperintensities had significantly shorter disease duration and greater disease severity, ie, a higher disease progression index, than those who did not. | | <https://pubmed.ncbi.nlm.nih.gov/7848130/> |
| **33** | | Cholinesterase inhibitors provide modest symptomatic benefit in mild–moderate AD. | These results indicate that cholinesterase inhibitors have a modest beneficial impact on neuropsychiatric and functional outcomes for patients with AD. | | <https://pubmed.ncbi.nlm.nih.gov/12517232/> |
|  |  |  | : Treatment with memantine/ChEI combination therapy in moderate-to-severe AD produces consistent benefits that appear to increase over time, and that are beyond those of ChEI treatment alone. | | <https://pubmed.ncbi.nlm.nih.gov/23110864/> |
|  |  |  | Cognitive function, as measured by the ADAS-cog, was significantly improved in the 5- and 10-mg/d donepezil groups as compared with the placebo group at weeks 12, 18, and 24. Clinician's global ratings on the CIBIC plus also improved in both the 5- and 10-mg/d donepezil groups relative to placebo. At the end of the 6-week placebo washout phase, ADAS-cog scores and CIBIC plus ratings were not significantly different for the three groups. Significant treatment benefits were also observed consistently in both the 5- and 10-mg/d groups on the MMSE and the CDR-SB, but there was no consistent effect on the patient-rated QoL. | | <https://pubmed.ncbi.nlm.nih.gov/9443470/> |
| **34** | | Memantine shows efficacy in moderate–severe AD. | Memantine showed a significant improvement in CF [standardized mean difference (SMD) = -0.24, 95% confidence intervals (95% CIs) = -0.34, -0.15, p < 0.00001, I2 = 35% ] and BD (SMD = -0.16, 95% CIs = -0.29, -0.04, p = 0.01, I2 = 52%) compared with placebo. In the sensitivity analysis including only patients with moderate-severe AD, memantine was superior to the placebo in reducing BD without considerable heterogeneity (SMD = -0.20, 95% CIs = -0.34, -0.07, p = 0.003, I2 = 36%). | | <https://pubmed.ncbi.nlm.nih.gov/28922160/> |
|  |  |  | Patients receiving memantine had a better outcome than those receiving placebo, according to the results of the CIBIC-Plus (P=0.06 with the last observation carried forward, P=0.03 for observed cases), the ADCS-ADLsev (P=0.02 with the last observation carried forward, P=0.003 for observed cases), and the Severe Impairment Battery (P<0.001 with the last observation carried forward, P=0.002 for observed cases). Memantine was not associated with a significant frequency of adverse events. | | <https://pubmed.ncbi.nlm.nih.gov/12672860/> |
|  |  |  | The change in total mean (SE) scores favored memantine vs placebo treatment for SIB (possible score range, 0-100), 0.9 (0.67) vs -2.5 (0.69), respectively (P<.001); ADCS-ADL19 (possible score range, 0-54), -2.0 (0.50) vs -3.4 (0.51), respectively (P =.03); and the CIBIC-Plus (possible score range, 1-7), 4.41 (0.074) vs 4.66 (0.075), respectively (P =.03). All other secondary measures showed significant benefits of memantine treatment. Treatment discontinuations because of adverse events for memantine vs placebo were 15 (7.4%) vs 25 (12.4%), respectively. | | <https://pubmed.ncbi.nlm.nih.gov/14734594/> |
| **35** | | Antipsychotics increase mortality risk in dementia. | Compared with respective matched nonusers, individuals receiving haloperidol had an increased mortality risk of 3.8% (95% CI, 1.0%-6.6%; P < .01) | | <https://pubmed.ncbi.nlm.nih.gov/25786075/> |
|  |  |  | Atypical antipsychotic drugs may be associated with a small increased risk for death compared with placebo. | | <https://pubmed.ncbi.nlm.nih.gov/16234500/> |
|  |  |  | Conventional antipsychotic medications were associated with a significantly higher adjusted risk of death than were atypical antipsychotic medications at all intervals studied (< or =180 days: relative risk, 1.37; 95 percent confidence interval, 1.27 to 1.49; <40 days: relative risk, 1.56; 95 percent confidence interval, 1.37 to 1.78; 40 to 79 days: relative risk, 1.37; 95 percent confidence interval, 1.19 to 1.59; and 80 to 180 days: relative risk, 1.27; 95 percent confidence interval, 1.14 to 1.41) and in all subgroups defined according to the presence or absence of dementia or nursing home residency. | | <https://pubmed.ncbi.nlm.nih.gov/16319382/> |
| **36** | | Statin use lowers risk of dementia | Statin use linked to lower risks of all-dementia, AD, and VaD. Numerous significant subgroup results highlight statins' diverse neuroprotective effects. | | <https://pubmed.ncbi.nlm.nih.gov/39822593/> |
|  |  |  | In the pooled analyses, statins were associated with a decreased risk of dementia [36 studies, OR 0.80 (CI 0.75-0.86)] and of AD [21 studies, OR 0.68 (CI 0.56-0.81)]. | | <https://pubmed.ncbi.nlm.nih.gov/34871380/> |
|  |  |  | Patients with statin had a lower all-caused dementia risk than those without statin (risk ratio [RR] 0.83, 95% CI 0.79-0.87, I2 = 57.73%). The overall pooled reduction of Alzheimer disease in patients with statin use was RR 0.69 (95% CI 0.60-0.80, p < 0.0001), and the overall pooled RR of statin use and vascular dementia risk was RR 0.93 (95% CI 0.74-1.16, p = 0.54). | | <https://pubmed.ncbi.nlm.nih.gov/31574510/> |
| **37** | | Antihypertensive therapy reduces risk of MCI. | Diuretic, ARB, and ACE-I use was, in addition to and/or independently of mean systolic blood pressure, associated with reduced risk of AD dementia in participants with normal cognition, while only diuretic use was associated with reduced risk in participants with MCI. | | <https://pubmed.ncbi.nlm.nih.gov/23911756/> |
|  |  |  | ARB users experienced a slower yearly global cognitive [2.5% of a SD, 95% CI = (0.1, 4.9)] and language [4.4% of a SD, 95% CI = (1.4, 7.4)] decline compared to non-users. The fully adjusted model reproduced similar associations for both global cognitive [β= 0.027, 95% CI = (-0.003, 0.057)], and language decline [β= 0.063, 95% CI = (0.023, 0.104)]. | | <https://pubmed.ncbi.nlm.nih.gov/35912747/> |
|  |  |  | Individuals treated with ARBs showed larger hippocampal volumes (R2 = 0.83, p = 0.05) and brain parenchymal fraction (R2 = 0.83, p = 0.01) than those treated with ACEIs. When stratified by diagnosis, this effect remained only in normal elderly adults and MCI patients, and a significant association between ARBs and lower WMH volume (R2 = 0.83, p = 0.03) emerged for AD patients only. | | <https://pubmed.ncbi.nlm.nih.gov/28731439/> |
| **38** | | NSAIDs have shown prevention benefit for AD. | In observational studies, use of non-steroidal anti-inflammatory drugs (NSAIDs) was significantly associated with a reduced risk of AD (RR, 0.72; 95%CI, 0.62-0.84) compared to no use of NSAIDs, especially in long term users (RR, 0.36; 95%CI, 0.17-0.74); the risks of AD were also lower in both aspirin (RR, 0.77; 95%CI, 0.63-0.95) and non-aspirin NSAID users (RR, 0.65; 95%CI, 0.47-0.88) compared with nonusers; whereas the use of corticosteroids showed no significant association (RR, 0.62; 95%CI, 0.26-1.46). In the single randomized controlled trial (RCT), NSAID use showed no significant effect on AD risk among dementia-free individuals (p > 0.05). | | <https://pubmed.ncbi.nlm.nih.gov/25227314/> |
|  |  |  | In this pooled dataset, nonsteroidal anti-inflammatory drug (NSAID) use reduced the risk of Alzheimer dementia (AD). | | <https://pubmed.ncbi.nlm.nih.gov/18509093/> |
|  |  |  | Results were consistent with previous cohort studies showing reduced risk of AD in NSAID users, but this association was found only in those with an APOE epsilon 4 allele, and there was no advantage for A beta(42)-lowering NSAIDs. | | <https://pubmed.ncbi.nlm.nih.gov/18003940/> |
| **39** | | Episodic memory impairment is the earliest hallmark in typical AD. | These results indicate that Alzheimer's disease is characterized by a long preclinical period during which episodic memory deficits are detectable. The magnitude of these deficits appears to be quite stable, at least up to 3 years before diagnosis. | | <https://pubmed.ncbi.nlm.nih.gov/11133790/> |
|  |  |  | These findings suggest that, when episodic memory is mildly impaired, limbic functions are still sufficient to subserve the remaining performance, whereas with more severe memory deficit resulting from accumulated pathology the neocortical areas that are normally involved in semantic memory are recruited, perhaps as a form of (inadequate) compensatory mechanism. | | <https://pubmed.ncbi.nlm.nih.gov/11960900/> |
|  |  |  | These results are consistent with a model in which Abeta deposition, hippocampal atrophy, and EM occur sequentially in elderly subjects, with Abeta deposition as the primary event in this cascade. This pattern suggests that declining EM in older individuals may be caused by Abeta-induced hippocampus atrophy. | | <https://pubmed.ncbi.nlm.nih.gov/19042931/> |
| **40** | | Semantic memory impairments are prominent in temporal-variant dementias. | In all patients, first symptoms involved semantics (4/6 LTLV, 1/6 RTLV), behavior (4/6 RTLV, 1/6 LTLV), or both (1 LTLV, 1 RTLV). Semantic loss began with anomia, word-finding difficulties, and repetitive speech, whereas the early behavioral syndrome was characterized by emotional distance, irritability, and disruption of physiologic drives (sleep, appetite, libido). | | <https://pubmed.ncbi.nlm.nih.gov/15851728/> |
|  |  |  | Subjects with tv-FTD showed severe deficits in semantic memory with preservation of attention and executive function. | | <https://pubmed.ncbi.nlm.nih.gov/10881252/> |
|  |  |  | Right anterior temporal lobe-predominant degeneration is characterized by early loss of empathy and person-specific knowledge, deficits that are caused by progressive decline in semantic memory for concepts of socioemotional relevance. | | <https://pubmed.ncbi.nlm.nih.gov/35731122/> |
| **41** | | Executive dysfunction is more pronounced in vascular and subcortical dementias. | Executive deficits are the most prominent cognitive characteristic associated with SIVD. | | <https://pubmed.ncbi.nlm.nih.gov/16361588/> |
|  |  |  | Subcortical ischaemic vascular disease is associated with subtle declines in executive functioning and visual memory, even in non-demented patients | | <https://pubmed.ncbi.nlm.nih.gov/11796772/> |
|  |  |  | The Alzheimer patients were more impaired than those with vascular dementia on episodic memory, while the patients with vascular dementia were more impaired on semantic memory, executive/attentional functioning, and visuospatial and perceptual skills. | | <https://pubmed.ncbi.nlm.nih.gov/14707310/> |
| **42** | | Visuospatial deficits are salient in DLB. | Visual perception is defective in probable DLB. The defective visual perception plays a role in development of visual hallucinations, delusional misidentifications, visual agnosias, and visuoconstructive disability charcteristic of DLB. | | <https://pubmed.ncbi.nlm.nih.gov/10768622/> |
|  |  |  | We conclude that the best model for differentiating DLB from Alzheimer's disease in the earliest stages of disease includes VH and visuospatial/constructional dysfunction, but not spontaneous EPS, as predictors. This suggests that clinical history plus a brief assessment of visuospatial function may be of the greatest value in correctly identifying DLB early during the course of disease. | | <https://pubmed.ncbi.nlm.nih.gov/16401618/> |
|  |  |  | Patients with LB pathology performed worse on the visuospatial dimension. | | <https://pubmed.ncbi.nlm.nih.gov/16247050/> |
| **43** | | Language deficits (anomia) are early in AD and FTD variants. | This study suggests the development of early anomia in MAPT mutation carriers, likely to be associated with impaired semantic knowledge. Clinical trials focused on the prodromal period within individuals with MAPT mutations should use language tasks, such as the BNT for patient stratification and as outcome measures. | | <https://pubmed.ncbi.nlm.nih.gov/35348856/> |
|  |  |  | We found significant naming deficits in all patient groups. | | <https://pubmed.ncbi.nlm.nih.gov/14761903/> |
|  |  |  | All cognitive domains showed substantial effect sizes, indicating cognitive impairment in bv-FTD patients compared to healthy controls. The cognitive domains with the largest effect sizes were social cognition, verbal memory and fluency (1.77–1.53). The cognitive profiles of bv-FTD and ALS … showed similarities on visual comparison and a moderate correlation … The cognitive profile of bv-FTD consists of deficits in social cognition, verbal memory, fluency and executive functions and shows similarities with the cognitive profile of ALS. These findings support a cognitive continuum encompassing ALS and bv-FTD. | | <https://pubmed.ncbi.nlm.nih.gov/29439163/> |
| **44** | | Working memory deficits appear in MCI and predict conversion. | he acceleration in cognitive decline occurred slightly earlier for semantic memory (76 months before diagnosis) and working memory (75 months) than other cognitive functions. Mild cognitive impairment was also preceded by years of cognitive decline that began earlier (80 months before diagnosis) and proceeded more rapidly (annual loss of 0.102 unit) in the amnestic than in the nonamnestic (62 months, 0.072 unit) subtype. | | <https://pubmed.ncbi.nlm.nih.gov/21403020/> |
|  |  |  | Mild cognitively impaired patients with memory plus other cognitive domain deficits, rather than those with pure amnestic MCI, constituted the high-risk group. Deficits in verbal memory and psychomotor speed/executive function abilities strongly predicted conversion to AD. | | <https://pubmed.ncbi.nlm.nih.gov/16894068/> |
|  |  |  | The activation differences between groups may be compensatory mechanisms within the MCI group for the effects of the putative AD neuropathology. This has been the first study that has examined verbal working memory in MCI. | | <https://pubmed.ncbi.nlm.nih.gov/20413893/> |
| **45** | | Inhibition deficits are common in AD. | Compared with the typical AD group, the frontal AD group performed significantly worse on 2 tests of frontal lobe functioning and on the Wechsler Adult Intelligence Scale-Revised Block Design test. No significant group differences were found on other tests. Analysis of brain tissue samples demonstrated that, despite comparable entorhinal, temporal, and parietal NFT loads, the frontal AD group showed a significantly higher NFT load in the frontal cortex than the typical AD group. | | <https://pubmed.ncbi.nlm.nih.gov/10520939/> |
|  |  |  | It was found that whilst most inhibitory mechanisms are affected by the disorder, some are relatively preserved, suggesting that inhibitory deficits in Alzheimer's disease may not be the result of a general inhibitory breakdown. In particular, the experimental results reviewed showed that Alzheimer's disease has a strong effect on tasks requiring controlled inhibition processes, such as the Stroop task. However, the presence of the disease appears to have relatively little effect on tasks requiring more automatic inhibition, such as the inhibition of return task. | | <https://pubmed.ncbi.nlm.nih.gov/14645147/> |
|  |  |  | Under these conditions, there is a very clear tendency for dual task performance to deteriorate while single task performance is maintained. | | <https://pubmed.ncbi.nlm.nih.gov/1782529/> |
| **46** | | Greater medial temporal atrophy is typical in dementia cases. | ] More than half (59.8%) of the patients had considerable MTA. Multiple linear regression analyses revealed that after correction for sex, age, education, and duration of dementia, neuropsychological tests showed that patients with higher grades of MTA or large vessel VaD had significantly worse general cognitive and executive functioning, whereas associations with small vessel disease were restricted to worse executive functioning. | | <https://pubmed.ncbi.nlm.nih.gov/17962598/> |
|  |  |  | MTA was more frequent and severe in all dementia groups compared with control subjects (AD, 100%; VaD, 88%; DLB, 62%; control subjects, 4%; p < 0.001). | | <https://pubmed.ncbi.nlm.nih.gov/10214736/> |
|  |  |  | The volume of each MTL structure was significantly smaller in AD patients than control subjects (p < 0.001). | | <https://pubmed.ncbi.nlm.nih.gov/9305341/> |
| **47** | | Frontal/insular atrophy is characteristic in bvFTD. | Rascovsky et al., 2011"group studies indicating that disproportionate frontal, insular or anterior temporal lobe atrophy (or combination thereof) may help distinguish bvFTD from healthy individuals, non-progressive behavioural syndromes and other dementias" | | <https://www.ncbi.nlm.nih.gov/pmc/articles/PMC3170532/> |
|  |  |  | The VBM studies in clinical variants confirmed established patterns of atrophy (SEMD, rostral temporal; bvFTD, mesial frontal; PNFA, left insula). FTD-U and FTD-T VBM results were very similar, showing severe atrophy in the temporal poles, mesial frontal lobe, and insulae. A conjunction analysis confirmed this similarity. | | <https://pubmed.ncbi.nlm.nih.gov/19433738/> |
|  |  |  | Patients with a CDR score of 0.5 had gray matter loss in frontal paralimbic cortices, but atrophy also involved a network of anterior cortical and subcortical regions. A CDR score of 1 showed more extensive frontal gray matter atrophy and white matter losses in corpus callosum and brainstem. A CDR score of 2 to 3 showed additional posterior insula, hippocampus, and parietal involvement, with white matter atrophy in presumed frontal projection fibers. | | Patients with a CDR score of 0.5 had gray matter loss in frontal paralimbic cortices, but atrophy also involved a network of anterior cortical and subcortical regions. A CDR score of 1 showed more extensive frontal gray matter atrophy and white matter losses in corpus callosum and brainstem. A CDR score of 2 to 3 showed additional posterior insula, hippocampus, and parietal involvement, with white matter atrophy in presumed frontal projection fibers. |
| **48** | | Occipital hypometabolism is common in DLB on FDG-PET. | Leonardo Iaccarino et al 2015 "SPM-t maps confirmed the typical temporal-parietal hypometabolic pattern with bilateral occipital hypometabolism involving also the primary visual cortex," | | <https://www.neurology.org/doi/10.1212/WNL.84.14_supplement.P2.182> |
|  |  |  | Positron emission tomographic studies have been reported to show occipital hypometabolism in dementia with Lewy bodies, in addition to the characteristic posterior bitemporal biparietal pattern of Alzheimer disease. | | <https://pubmed.ncbi.nlm.nih.gov/11255457/> |
|  |  |  | In summary, a sizeable group of clinically diagnosed ADD and aMCI patients exhibit posterior-occipital FDG-PET patterns typically associated with Lewy body pathology, and these also show less abnormal Alzheimer's disease biomarkers as well as specific clinical features typically associated with dementia with Lewy bodies. | | <https://pubmed.ncbi.nlm.nih.gov/37284793/> |
| **49** | | Posterior cingulate hypometabolism occurs early in AD. | Hippocampal atrophy and hypometabolism of the posterior cingulate cortex (PCC), early markers of Alzheimer's disease (AD), have been shown to be associated in late mild cognitive impairment and early AD via atrophy of connecting cingulum fibers. | | <https://pubmed.ncbi.nlm.nih.gov/29036815/> |
|  |  | Prediction and analysis of actual patients consistently indicated marked metabolic reduction (21-22%) in the posterior cingulate cortex and cinguloparietal transitional area in patients with very early Alzheimer's disease. Mean metabolic reduction in the posterior cingulate cortex was significantly greater than that in the lateral neocortices or parahippocampal cortex. The result suggests a functional importance for the posterior cingulate cortex in impairment of learning and memory, which is a feature of very early Alzheimer's disease. | | <https://pubmed.ncbi.nlm.nih.gov/9225689/> | |
|  |  | PMCI had a significantly decreased rCBF in the left posterior cingulate cortex, as compared to SMCI. Left posterior cingulate rCBF ratios were entered into a logistic regression model for ROC curve calculation. The area under the ROC curve was 74%-76%, which indicates an acceptable discrimination between PMCI and SMCI at baseline. | | <https://pubmed.ncbi.nlm.nih.gov/12227833/> | |
| **50** | | Hearing loss is associated with an increased risk of developing dementia. | Having a hearing loss was associated with an increased risk of dementia, with an adjusted hazard ratio (HR) of 1.07 (95% CI, 1.04-1.11) compared with having no hearing loss. | | <https://pubmed.ncbi.nlm.nih.gov/38175662/> |
|  |  |  | Hearing loss had a negative association with cognition; for those with moderate to severe loss, the score on memory assessment was a full 1 point less (-1.00; 95% CI, -1.24 to -0.76), ceteris paribus, relative to those with no hearing loss. However, this association was seen only in the individuals with untreated hearing loss (ie, those who did not use hearing aids) (-1.16; 95% CI, -1.45 to -0.87). | | <https://pubmed.ncbi.nlm.nih.gov/30193368/> |
|  |  |  | Adjusting for numerous confounders, an increased risk of disability and dementia was found for participants reporting hearing problems. | | <https://pubmed.ncbi.nlm.nih.gov/29304204/> |
| **51** | | Social isolation increases risk of dementia. | Loneliness increased risk for all-cause dementia (HR = 1.306, 95% CI [1.197,1.426]), Alzheimer's disease (HR = 1.393, 95% CI [1.290,1.504]; k = 5), vascular dementia (HR = 1.735, 95% CI [1.483,2.029]; k = 3), and cognitive impairment (HR = 1.150, 95% CI [1.113,1.189]). | | <https://pubmed.ncbi.nlm.nih.gov/39802418/> |
|  |  |  | Cox proportional hazards regression indicated that loneliness was associated with a 40% increased risk of dementia. | | <https://pubmed.ncbi.nlm.nih.gov/30365023/> |
|  |  |  | Low social participation (RR: 1.41 (95% CI: 1.13-1.75)), less frequent social contact (RR: 1.57 (95% CI: 1.32-1.85)), and more loneliness (RR: 1.58 (95% CI: 1.19-2.09)) were statistically significant associated with incident dementia. | | <https://pubmed.ncbi.nlm.nih.gov/25956016/> |
| **52** | | Poor sleep quality is associated with higher amyloid burden. | Self-reported shorter sleep duration was linearly associated with higher Aβ burden (β [SE] = -0.01 [0.00]; P = .005), and short sleep duration was associated with reduced cognition that was mostly in memory domains. No difference in Aβ was found between long and normal sleep duration groups (β [SE] = 0.00 [0.01]; P = .99). | | <https://pubmed.ncbi.nlm.nih.gov/34459862/> |
|  |  |  | Worse subjective sleep quality, more sleep problems, and daytime somnolence were associated with greater AD pathology, indicated by lower CSF Aβ42/Aβ40 and higher t-tau/Aβ42, p-tau/Aβ42, MCP-1/Aβ42, and YKL-40/Aβ42. There were no significant associations between sleep and NFL or neurogranin. | | <https://pubmed.ncbi.nlm.nih.gov/28679595/> |
|  |  |  | Daytime sleepiness was associated with Aβ deposition in the brainstem (B = 0.0063; 95% CI, 0.001 to 0.012; P = .02), but not MMSE performance (B = -0.01; 95% CI, -0.39 to 0.37; P = .96). The number of nocturnal awakenings was associated with Aβ deposition in the precuneus (B = 0.11; 95% CI, 0.06 to 0.17; P < .001) and poor MMSE performance (B = -2.13; 95% CI, -3.13 to -1.13; P < .001). Mediation analysis demonstrated an indirect association between Aβ deposition and poor MMSE performance that relied on nocturnal awakenings as an intermediary (B = -3.99; 95% CI, -7.88 to -0.83; P = .01). | | <https://pubmed.ncbi.nlm.nih.gov/31617927/> |
| **53** | | Physical activity is associated with reduced risk of cognitive impairment. | PA was associated with a decreased risk of all-cause dementia (pooled relative risk 0.80, 95% CI 0.77 to 0.84, n=257 983), Alzheimer’s disease (0.86, 95% CI 0.80 to 0.93, n=128 261) and vascular dementia (0.79, 95% CI 0.66 to 0.95, n=33 870), even in longer follow-ups (≥20 years) for all-cause dementia and Alzheimer’s disease. | | <https://pubmed.ncbi.nlm.nih.gov/35301183/> |
|  |  |  | Meta-analysis, using the quality-effects model, suggests that participants with higher levels of physical activity, when compared to those with lower levels, are at reduced risk of cognitive decline, RR 0.65, 95% CI 0.55-0.76, and dementia, RR 0.86, 95% CI 0.76-0.97. | | <https://pubmed.ncbi.nlm.nih.gov/24885250/> |
|  |  |  | High levels of physical activity were associated with reduced risks of cognitive impairment (age-, sex-, and education-adjusted odds ratio, 0.58; 95% confidence interval, 0.41-0.83), Alzheimer disease (odds ratio, 0.50; 95% confidence interval, 0.28-0.90), and dementia of any type (odds ratio, 0.63; 95% confidence interval, 0.40-0.98). | | <https://pubmed.ncbi.nlm.nih.gov/11255456/> |
| **54** | | HPA-axis dysregulation (cortisol) associates with hippocampal atrophy. | Here we demonstrate that aged humans with significant prolonged cortisol elevations showed reduced hippocampal volume and deficits in hippocampus-dependent memory tasks compared to normal-cortisol controls. | | <https://pubmed.ncbi.nlm.nih.gov/10195112/> |
|  |  |  | Plasma cortisol predicted greater hippocampal atrophy, such that participants with higher cortisol showed faster decline in hippocampal volume over time (interaction: β = -0.15, p = 0.004). Small hippocampal volume predicted a higher risk of clinical progression to AD (haard ratio [HR] = 2.15; confidence in terval [CI], 1.64-2.80; p < 0.001). | | <https://pubmed.ncbi.nlm.nih.gov/37583892/> |
|  |  |  | Further, basal plasma cortisol levels were correlated with the radial width of the temporal horn, and elevated levels of plasma cortisol predicted a worse general cognitive performance. Higher plasma cortisol levels also correlated with rapid declines in MMSE scores after 2 years. | | <https://pubmed.ncbi.nlm.nih.gov/19570680/> |
| **55** | | Donepezil improves cognition in MCI and mild–moderate AD. | Donepezil hydrochloride (5 and 10 mg) administered once daily is a well-tolerated and efficacious agent for treating the symptoms of mild to moderately severe Alzheimer disease. | | <https://pubmed.ncbi.nlm.nih.gov/9588436/> |
|  |  |  | In patients with moderate or severe Alzheimer's disease, continued treatment with donepezil was associated with cognitive benefits that exceeded the minimum clinically important difference and with significant functional benefits over the course of 12 months. | | <https://pubmed.ncbi.nlm.nih.gov/22397651/> |
|  |  |  | Perceived Deficits Questionnaire scores favored donepezil at week 24 (p = 0.05). Clinical Global Impression of Change-MCI scores favored donepezil only at week 6 (p = 0.04). | | <https://pubmed.ncbi.nlm.nih.gov/19176895/> |
| **56** | | Galantamine improves cognition in mild–moderate AD. | At six months, patients who received galantamine had a significantly better outcome on the 11 item cognitive subscale of the Alzheimer's disease assessment scale than patients in the placebo group (mean treatment effect 2.9 points for lower dose and 3.1 for higher dose, intention to treat analysis, P<0.001 for both doses). | | <https://pubmed.ncbi.nlm.nih.gov/11110737/> |
|  |  |  | Galantamine significantly improved cognitive function relative to placebo; the treatment effects were 3.9 points (lower dose) and 3.8 points (higher dose) on the ADAS-cog/11 scale at month 6 (p < 0.001 in both cases). Both doses of galantamine produced a better outcome on CIBIC-plus than placebo (p < 0.05). | | <https://pubmed.ncbi.nlm.nih.gov/10881250/> |
|  |  |  | Mean CDR-sum of boxes declined less with galantamine than placebo at 12 and 24 months in Study 1 (p = 0.024 [12 months] and p = 0.028 [24 months]), but not in Study 2 (p = 0.662 [12 months] and p = 0.056 [24 months]). Digit Symbol Substitution Test scores improved with galantamine in Study 1 at 12 months and in Study 2 at 24 months (Study 1: p = 0.009 [month 12] and p = 0.079 [Month 24]; Study 2: p = 0.154 [month 12] and p = 0.020 [month 24]). | | <https://pubmed.ncbi.nlm.nih.gov/18322263/> |
| **57** | | Combined donepezil+memantine yields additive benefits in moderate–severe AD. | In patients with moderate or severe Alzheimer's disease, continued treatment with donepezil was associated with cognitive benefits that exceeded the minimum clinically important difference and with significant functional benefits over the course of 12 months. | | <https://pubmed.ncbi.nlm.nih.gov/22397651/> |
|  |  |  | Here, we show that the combined use of Donepezil and Memantine significantly elevates the probability of five-year survival. In particular, their combined use increases the probability of five-year survival by 0.050 (0.021, 0.078) (6.4%), 0.049 (0.012, 0.085), (6.3%), 0.065 (0.035, 0.095) (8.3%) compared to no drug treatment, the Memantine monotherapy, and the Donepezil monotherapy respectively. We also identify a significant beneficial additive drug-drug interaction effect between Donepezil and Memantine of 0.064 (0.030, 0.098). | | <https://pubmed.ncbi.nlm.nih.gov/38783011/> |
|  |  |  | Frontal assessment battery (FAB) declined significantly at 12 months after memantine addition in the donepezil subgroup, while the galantamine subgroup significantly improved at 6 months. Affective functions were well preserved after memantine addition until 12 months, except for the apathy scale at 12 months after memantine addition in the galantamine subgroup. The combination therapy of donepezil plus memantine was better for apathy in older AD patients, and galantamine plus memantine was better for cognitive functions. | | <https://pubmed.ncbi.nlm.nih.gov/25624417/> |
| **58** | | Aspirin did not prevent cognitive decline in healthy older adults (ASPREE-Cog). | At baseline, mean vocabulary scores (an indicator of previous cognitive ability) were similar in the aspirin (30.9, SD 4.7) and placebo (31.1, SD 4.7) groups. In the primary intention to treat analysis, there was no significant difference at follow-up between the groups in the proportion achieving over the median general factor cognitive score (32.7% and 34.8% respectively, odds ratio 0.91, 95% confidence interval 0.79 to 1.05, P=0.20) or in mean scores on the individual cognitive tests. There were also no significant differences in change in cognitive ability over the five years in a subset of 504 who underwent detailed cognitive testing at baseline. | | <https://pubmed.ncbi.nlm.nih.gov/18762476/> |
|  |  |  | A total of 19,114 participants were followed over a median 4.7 years and 964 triggered further dementia assessments. There were 575 adjudicated dementia cases, and 41% were classified as clinically probable AD. There was no substantial difference in the risk of all dementia triggers (hazard ratio [HR], 1.03; 95% confidence interval [CI], 0.91-1.17), probable AD (HR, 0.96; 95% CI, 0.74-1.24), or MCI (HR, 1.12; 95% CI, 0.92-1.37) between aspirin and placebo. Cognitive change over time was similar in the aspirin and placebo groups. | | <https://pubmed.ncbi.nlm.nih.gov/32213642/> |
|  |  |  | At the initial assessment (mean 5.6 years after randomisation) cognitive performance in the aspirin group was similar to that of the placebo group (mean difference in global score -0.01, 95% confidence interval -0.04 to 0.02). Mean decline in the global score from the first to the final cognitive assessment was also similar in the aspirin compared with placebo groups (mean difference 0.01, -0.02 to 0.04). The risk of substantial decline (in the worst 10th centile of decline) was also comparable between the groups (relative risk 0.92, 0.77 to 1.10). Findings were similar for verbal memory; however, a 20% lower risk was observed for decline in category fluency with aspirin (relative risk 0.80, 0.67 to 0.97). | | <https://pubmed.ncbi.nlm.nih.gov/17468120/> |
| **59** | | Amyloid-positive cognitively normal adults decline faster than amyloid-negative peers. | Among the 445 participants (243 with normal amyloid, 202 with elevated amyloid), mean (SD) age was 74.0 (5.9) years, mean education was 16.4 (2.7) years, and 52% were women. The mean score for PACC at baseline was 0.00 (2.60); for MMSE, 29.0 (1.2); for CDR-Sum of Boxes, 0.04 (0.14); and for Logical Memory Delayed Recall, 13.1 (3.3). Compared with the group with normal amyloid, those with elevated amyloid had worse mean scores at 4 years on the PACC (mean difference, 1.51 points [95% CI, 0.94-2.10]; P < .001), MMSE (mean difference, 0.56 points [95% CI, 0.32-0.80]; P < .001), and CDR-Sum of Boxes (mean difference, 0.23 points [95% CI, 0.08-0.38]; P = .002). For Logical Memory Delayed Recall, between-group score was not statistically significant at 4 years (mean difference, 0.73 story units [95% CI, -0.02 to 1.48]; P = .056). | | <https://pubmed.ncbi.nlm.nih.gov/28609533/> |
|  |  |  | Higher Abeta deposition is associated with greater longitudinal decline in mental status and verbal memory in the preceding years. | | <https://pubmed.ncbi.nlm.nih.gov/20147655/> |
|  |  |  | Linear mixed-model analyses adjusted for baseline cognitive function indicated that, relative to individuals with low cerebral Aβ, individuals with high cerebral Aβ showed significantly greater decline in working memory and verbal and visual episodic memory at 18 months. Compared with noncarriers, APOE ε4 carriers showed a greater decline in visual memory at the 18-month assessment. No interaction between APOE ε4 and cerebral Aβ load was observed for any measure of cognitive function. | | <https://pubmed.ncbi.nlm.nih.gov/23071163/> |
| **60** | | CSF biomarkers become abnormal years before clinical symptoms. | Concentrations of amyloid-beta (Aβ)42 in the CSF appeared to decline 25 years before expected symptom onset. Aβ deposition, as measured by positron-emission tomography with the use of Pittsburgh compound B, was detected 15 years before expected symptom onset. Increased concentrations of tau protein in the CSF and an increase in brain atrophy were detected 15 years before expected symptom onset. | | <https://pubmed.ncbi.nlm.nih.gov/22784036/> |
|  |  |  | Importantly, CSF tau/Aβ42 ratios show strong promise as antecedent (preclinical) biomarkers that predict future dementia in cognitively normal older adults. | | <https://pubmed.ncbi.nlm.nih.gov/17210801/> |
|  |  |  | MRI and CSF provide complimentary predictive information about time to conversion from amnestic mild cognitive impairment to Alzheimer disease and combination of the 2 provides better prediction than either source alone. | | <https://pubmed.ncbi.nlm.nih.gov/19636048/> |
| **61** | | Lower clock-drawing performance scores are associated with poorer functional abilities in individuals with dementia. | The DCR scaled well with FAQ scores, and ML classifiers trained on multimodal DCR features demonstrated strong performance in predicting functional impairment on a held-out test set. Differences in FAQ scores between DCR-predicted classes were comparable across key demographic groups. | | <https://pubmed.ncbi.nlm.nih.gov/39533697/> |
|  |  |  | The optimal model (500 features) classified functional impairment with 0.77 sensitivity, 0.82 specificity, 0.90 negative predictive value (NPV), 0.61 positive predictive value (PPV), 0.86 area under the receiver operating characteristic curve (AUC), and 0.80 accuracy. The most predictive features were the duration of speech during recall and the placement of clock components. | | <https://pmc.ncbi.nlm.nih.gov/articles/PMC11712186/> |
|  |  |  | LHQ significantly predicted mFD and modFD (p’s<0.001, R2 = 0.13, and R2 = 0.12). Adding DCR metrics to the model improved both models (p’s<0.001, R2’s = 0.18 and 0.13, respectively). Surviving LHQ questions for predicting mFD related to unintended weight loss, social engagement/volunteering, cognitively stimulating tasks, sleep, and life satisfaction/purpose. Surviving LHQ questions for predicting modFD related to unintended weight loss, social engagement/volunteering, cognitively stimulating tasks, and sleep. | | <https://alz-journals.onlinelibrary.wiley.com/doi/10.1002/alz.083113> |
| **62** | | Atrial fibrillation is associated with higher risk of cognitive impairment. | Atrial fibrillation was significantly associated with a higher risk for cognitive impairment in patients with first-ever or recurrent stroke (relative risk [RR], 2.70 [95% CI, 1.82 to 4.00]) and in a broader population including patients with or without a history of stroke (RR, 1.40 [CI, 1.19 to 1.64]). | | <https://pubmed.ncbi.nlm.nih.gov/23460057/> |
|  |  |  | Compared with AF-free participants, those with longer exposure to AF (5, 10, or 15 years) experienced faster cognitive decline after adjustment for sociodemographic, behavioural, and chronic diseases (P for trend = 0.01). | | <https://pubmed.ncbi.nlm.nih.gov/28460139/> |
|  |  |  | In addition, incident AF was associated with an increased risk of dementia (hazard ratio, 1.23; 95% confidence interval, 1.04-1.45), after adjusting for cardiovascular risk factors, including ischemic stroke. | | <https://pubmed.ncbi.nlm.nih.gov/29514809/> |
| **63** | | Chronic kidney disease is associated with cognitive impairment. | Meta-analysis of cross-sectional and longitudinal studies comprising 54,779 participants yielded an association of cognitive decline in patients with CKD compared with patients without CKD (OR 1.65, 95% CI 1.32-2.05; p < 0.001, and OR 1.39, 95% CI 1.15-1.68; p < 0.001, respectively). | | <https://pubmed.ncbi.nlm.nih.gov/22555151/> |
|  |  |  | CKD patients (eGFR < 60 mL/min/1.73 m2) performed worse than control groups (eGFR ≥ 60 mL/min/1.73 m2) on Orientation & Attention (SMD -0.79, 95% CI, -1.44 to -0.13), Language (SMD -0.63, 95% CI, -0.85 to -0.41), Concept Formation & Reasoning (SMD -0.63, 95% CI, -1.07 to -0.18), Executive Function (SMD -0.53, 95% CI, -0.85 to -0.21), Memory (SMD -0.48, 95% CI, -0.79 to -0.18), and Global Cognition (SMD -0.48, 95% CI, -0.72 to -0.24). | | <https://pubmed.ncbi.nlm.nih.gov/27964726/> |
|  |  |  | People with CKD have a high prevalence of CI, especially in patients with hemodialysis. | | <https://pubmed.ncbi.nlm.nih.gov/38829896/> |
| **64** | | Hippocampal atrophy rate is faster in AD than normal aging. | The annualized rates of hippocampal volume loss for each of the three initial clinical groups decreased progressively in the following order: AD > MC > C. Within the control and MCI groups, those who declined had a significantly greater rate of volume loss than those who remained clinically stable. The mean annualized rates of hippocampal atrophy by followup clinical group were: control-stable 1.73%, control-decliner 2.81%, MCI-stable 2.55%, MCI-decliner 3.69 %, AD 3.5% | | <https://pmc.ncbi.nlm.nih.gov/articles/PMC2724764/> |
|  |  |  | The median rate of atrophy was significantly greater in the Alzheimer's disease group than in the control group (12·3 [range 5·8 to 23·6] vs 0·3 (−1·2 to 1·7) mL per year; p<0·0001). There was no overlap between the groups. Furthermore, three non-demented individuals at risk of familial Alzheimer's disease had scans 6–14 months apart and showed greater rates of volume loss than the controls; these three individuals have subsequently developed symptoms. | | <https://www.thelancet.com/journals/lancet/article/PIIS0140-6736(96)05228-2/abstract> |
|  |  |  | Nine studies were included from seven centres, with data from a total of 595 AD and 212 matched controls. Mean (95% CIs) annualised hippocampal atrophy rates were found to be 4.66% (95% CI 3.92, 5.40) for AD subjects and 1.41% (0.52, 2.30) for controls. | | <https://pmc.ncbi.nlm.nih.gov/articles/PMC2773132/> |
| **65** | | Lower cortical thickness predicts faster cognitive decline. | In presymptomatic individuals harboring indicators of AD, baseline thickness in AD-vulnerable cortical regions was significantly reduced compared with that of healthy control individuals, but baseline hippocampal volume was not. | | <https://pubmed.ncbi.nlm.nih.gov/21825241/> |
|  |  |  | Thinning in specific cortical areas known to be affected by Alzheimer disease (AD) is detectable in individuals with questionable AD dementia (QAD) and predicts conversion to mild AD dementia. | | <https://pmc.ncbi.nlm.nih.gov/articles/PMC2677470/> |
|  |  |  | Baseline cortical thickness as well as hippocampal volume predicted cognitive decline, regardless of baseline cognitive status. In individuals unimpaired at baseline, decreases in cortical thickness and hippocampal volume independently predicted cognitive decline. For participants with baseline mild impairment, decreases in hippocampal volume predicted further cognitive decline. | | <https://pubmed.ncbi.nlm.nih.gov/40676867/> |
| **66** | | Blood levels of amyloid-β indicate brain amyloid plaque deposition. | All test biomarkers showed high performance when predicting brain amyloid-β burden. In particular, the composite biomarker showed very high areas under the receiver operating characteristic curves (AUCs) in both data sets (discovery, 96.7%, n = 121 and validation, 94.1%, n = 111) with an accuracy approximately equal to 90% when using PIB-PET as a standard of truth. Furthermore, test biomarkers were correlated with amyloid-β-PET burden and levels of Aβ1–42 in cerebrospinal fluid. | | <https://pubmed.ncbi.nlm.nih.gov/29420472/> |
|  |  |  | The TP42/40 plasma ratio was significantly reduced in amyloid-PET-positive participants at all time points (p < 0.0001). | | <https://pubmed.ncbi.nlm.nih.gov/32179698/> |
|  |  |  | In the combined cohort of 465 participants, plasma Aβ42/Aβ40 had good concordance with amyloid PET status (receiver operating characteristic area under the curve [AUC] 0.84, 95% confidence interval [CI] 0.80-0.87); concordance improved with the inclusion of APOE ε4 carrier status (AUC 0.88, 95% CI 0.85-0.91). | | <https://pubmed.ncbi.nlm.nih.gov/34906975/> |
| **67** | | Blood levels of p-tau217 distinguish Alzheimer’s disease from other neurodegenerative disorders. | Among 1402 participants from 3 selected cohorts, plasma P-tau217 discriminated AD from other neurodegenerative diseases, with significantly higher accuracy than established plasma- and MRI-based biomarkers, and its performance was not significantly different from key CSF- or PET-based measures. | | <https://pubmed.ncbi.nlm.nih.gov/32722745/> |
|  |  |  | Both p-tau217 and p-tau181 had excellent diagnostic performance for differentiating patients with Alzheimer's disease syndromes from other neurodegenerative disorders. There was some evidence in favour of p-tau217 compared with p-tau181 for differential diagnosis of Alzheimer's disease syndromes versus FTLD syndromes, as an indication of amyloid-PET-positivity, and for stronger correlations with tau-PET signal. | | <https://pmc.ncbi.nlm.nih.gov/articles/PMC8711249/> |
|  |  |  | Plasma p-tau217 effectively distinguished AD from FTLD, with the NfL/p-tau217 ratio showing superior accuracy. The three-range approach identified thresholds with 95% and 97.5% sensitivity and specificity, reducing the need for cerebrospinal fluid testing by 75% and 54%, respectively. | | <https://pubmed.ncbi.nlm.nih.gov/39776166/> |
| **68** | | Plasma neurofilament light chain levels correlate with future cognitive decline. | In this case-control study of 193 cognitively healthy controls, 197 patients with mild cognitive impairment, and 180 patients with Alzheimer disease dementia, plasma neurofilament light was associated with Alzheimer disease and correlated with future progression of cognitive decline, brain atrophy, and brain hypometabolism. | | <https://pmc.ncbi.nlm.nih.gov/articles/PMC5822204/> |
|  |  |  | Faster increase in NfL levels correlated with faster increase in CSF biomarkers of neuronal injury, faster rates of atrophy and hypometabolism, and faster worsening in global cognition (all P < .05 in patients with mild cognitive impairment; associations differed slightly in cognitively unimpaired controls and patients with AD dementia). | | <https://pubmed.ncbi.nlm.nih.gov/31009028/> |
|  |  |  | These observations support NfL in both CSF and blood as an early marker of neurodegeneration but suggest that NfL measured in the CSF may be better suited for monitoring clinical trial outcomes in symptomatic AD patients. | | <https://pmc.ncbi.nlm.nih.gov/articles/PMC11574007/> |
| **69** | | Plasma GFAP is elevated in amyloid-positive individuals. | We found that plasma GFAP concentration was significantly increased in all amyloid-β-positive groups compared with participants without amyloid-β pathology (P < 0.01). | | <https://pmc.ncbi.nlm.nih.gov/articles/PMC8677538/> |
|  |  |  | Plasma GFAP levels were significantly higher in individuals with preclinical AD in comparison with cognitively unimpaired (CU) Aβ-negative individuals (TRIAD: Aβ-negative mean [SD], 185.1 [93.5] pg/mL, Aβ-positive mean [SD], 285.0 [142.6] pg/mL; ALFA+: Aβ-negative mean [SD], 121.9 [42.4] pg/mL, Aβ-positive mean [SD], 169.9 [78.5] pg/mL). | | <https://pubmed.ncbi.nlm.nih.gov/34661615/> |
|  |  |  | Plasma GFAP levels were significantly higher (p < 0.00001), and plasma Aβ1–42/Aβ1–40 ratios were significantly lower (p < 0.005), in Aβ+ participants compared to Aβ− participants, adjusted for covariates age, sex, and apolipoprotein E-ε4 carriage. | | <https://pmc.ncbi.nlm.nih.gov/articles/PMC7801513/> |
| **70** | | Retinal changes are associated with the presence of Alzheimer’s disease. | n the internal validation dataset, the deep learning model had 83·6% (SD 2·5) accuracy, 93·2% (SD 2·2) sensitivity, 82·0% (SD 3·1) specificity, and an area under the receiver operating characteristic curve (AUROC) of 0·93 (0·01) for detecting Alzheimer's disease-dementia. In the testing datasets, the bilateral deep learning model had accuracies ranging from 79·6% (SD 15·5) to 92·1% (11·4) and AUROCs ranging from 0·73 (SD 0·24) to 0·91 (0·10). | | <https://pubmed.ncbi.nlm.nih.gov/36192349/> |
|  |  |  | Retinal thickness is decreased in AD and MCI patients compared to HC. This confirms that neurodegenerative diseases may be reflected by retinal changes. | | <https://pubmed.ncbi.nlm.nih.gov/28275698/> |
|  |  |  | The surface area of inclusion bodies increased as a function of cortical amyloid burden. Additionally, there was a trend toward a selective volume increase in the inner plexiform layer (IPL; a layer rich in cholinergic activity) of the retina in Aβ+ relative to Aβ- participants, and IPL volume was correlated with the surface area of retinal inclusion bodies. | | <https://pubmed.ncbi.nlm.nih.gov/27830174/> |
| **71** | | Hormone replacement therapy in postmenopausal women is associated with improved global cognition. | Results indicate that women who had used estrogen replacement scored significantly higher on cognitive testing at baseline than nonusers, and their performance on verbal memory improved slightly over time. The effect of estrogen on cognition was independent of age, education, ethnicity, and APOE genotype. | | <https://pubmed.ncbi.nlm.nih.gov/9484355/> |
|  |  |  | In a population cohort of older women, lifetime HRT exposure was associated with improved global cognition and attenuated decline over a 3-year interval. Improvements were greatest in the oldest old. | | <https://pubmed.ncbi.nlm.nih.gov/11756599/> |
|  |  |  | HRT introduction is associated with improved delayed memory and larger entorhinal and amygdala volumes in APOE4 carriers only. | | <https://pubmed.ncbi.nlm.nih.gov/36624497/> |
| **72** | | Hearing aid use mitigates accelerated cognitive decline from hearing loss. | Hearing impairment was associated with increased risk of MCI (standardized hazard ratio [HR] 2.58, 95% confidence interval [CI: 1.73 to 3.84], P = .004) and an accelerated rate of cognitive decline (P < .001). Hearing aid users were less likely to develop MCI than hearing‐impaired individuals who did not use a hearing aid (HR 0.47, 95% CI [0.29 to 0.74], P = .001). No difference in risk of MCI was observed between individuals with normal hearing and hearing‐impaired adults using hearing aids (HR 0.86, 95% CI [0.56 to 1.34], P = .51). | | <https://pmc.ncbi.nlm.nih.gov/articles/PMC8863441/> |
|  |  |  | Compared with people without hearing loss, the risk of dementia was higher among people with hearing loss who were not using hearing aids than those who had hearing loss and were using hearing aids, with HRs of 1.20 (95% CI, 1.13-1.27) and 1.06 (95% CI, 1.01-1.10), respectively. | | <https://pmc.ncbi.nlm.nih.gov/articles/PMC10767640/> |
|  |  |  | These findings suggest that a hearing intervention might reduce cognitive change over 3 years in populations of older adults at increased risk for cognitive decline but not in populations at decreased risk for cognitive decline. | | <https://pubmed.ncbi.nlm.nih.gov/37478886/> |
| **73** | | Stroop performance declines with AD severity. | The Stroop Color-Word Test (SCWT; C. Golden, 1978) was examined in 59 patients with probable Alzheimer's disease (AD) and in 51 demographically comparable normal control (NC) participants. AD patients produced significantly larger Stroop interference effects than NC participants, and level of dementia severity significantly influenced SCWT performance. | | <https://pubmed.ncbi.nlm.nih.gov/12146681/> |
|  |  |  | Slowing on color naming and word reading was observed, and was greater in moderate than in mild dementia subjects. | | <https://pubmed.ncbi.nlm.nih.gov/6480252/> |
|  |  |  | We found a significant increase in Stroop effects for AD patients, across studies. This AD-related change was associated with a slowing in SOP. However, after correcting for a bias in the distribution of latencies, SOP could only explain a moderate portion of the total variance (25%). Moreover, we found strong evidence for an AD-related increase in the latency difference between naming the font-color and reading color-neutral stimuli (r2 = 0.98). | | <https://pubmed.ncbi.nlm.nih.gov/24100125/> |
| **74** | | Cortical default mode hubs (PCC) show early amyloid deposition. | We show that Aβ accumulation preferentially starts in the precuneus, medial orbitofrontal, and posterior cingulate cortices, i.e., several of the core regions of the default mode network (DMN). This early pattern of Aβ accumulation is already evident in individuals with normal Aβ42 in the CSF and normal amyloid PET who subsequently convert to having abnormal CSF Aβ42. The earliest Aβ accumulation is further associated with hypoconnectivity within the DMN and between the DMN and the frontoparietal network, but not with brain atrophy or glucose hypometabolism. Our results suggest that Aβ fibrils start to accumulate predominantly within certain parts of the DMN in preclinical AD and already then affect brain connectivity. | | <https://pubmed.ncbi.nlm.nih.gov/29089479/> |
|  |  |  | Even below the PRC Aβ threshold, DMN function remains related to PRC Aβ deposition and cognitive performance. | | <https://pubmed.ncbi.nlm.nih.gov/39559982/> |
|  |  |  | Cortical accumulation of amyloid beta is one of the first events of Alzheimer's disease pathophysiology, and has been suggested to follow a consistent spatiotemporal ordering, starting in the posterior cingulate cortex, precuneus and medio-orbitofrontal cortex. | | <https://pubmed.ncbi.nlm.nih.gov/34617016/> |
| **75** | | Positive family history increases risk of dementia independent of APOE. | Parental history was associated with risk of dementia independently of known genetic risk factors (hazard ratio [HR] 1.67, 95% confidence interval [CI] 1.12–2.48), in particular when parents were diagnosed at younger age (<80 years: HR 2.58, 95% CI 1.61–4.15; ≥80 years: HR 1.01, 95% CI 0.58–1.77). | | <https://pubmed.ncbi.nlm.nih.gov/28356461/> |
|  |  |  | Parental family history of AD was associated with lower FA in regions of the brain known to be affected by AD, including cingulum, corpus callosum, tapetum, uncinate fasciculus, hippocampus, and adjacent white matter. Contrary to previous reports there was no main effect of ApoE ε4; however, there was an additive effect of family history and ApoE ε4 where family history positive participants who were also ApoE ε4 carriers had the lowest FA compared to the other groups. | | <https://pmc.ncbi.nlm.nih.gov/articles/PMC2933285/> |
|  |  |  | Our findings suggest that lower plasma apoE levels in middle age could be a risk factor for Alzheimer disease in old age, independent of APOE genotype. | | <https://pmc.ncbi.nlm.nih.gov/articles/PMC2734293/> |
| **76** | | Benzodiazepine use is linked to cognitive impairment. | Over a mean follow-up of 7.3 years, 797 participants (23.2%) developed dementia, of whom 637 developed Alzheimer's disease. For dementia, the adjusted hazard ratios associated with cumulative benzodiazepine use compared with non-use were 1.25 (95% confidence interval 1.03 to 1.51) for 1-30 TSDDs; 1.31 (1.00 to 1.71) for 31-120 TSDDs; and 1.07 (0.82 to 1.39) for ≥ 121 TSDDs. Results were similar for Alzheimer's disease. Higher benzodiazepine use was not associated with more rapid cognitive decline. | | <https://pubmed.ncbi.nlm.nih.gov/26837813/> |
|  |  |  | During a 15 year follow-up, 253 incident cases of dementia were confirmed. New use of benzodiazepines was associated with an increased risk of dementia (multivariable adjusted hazard ratio 1.60, 95% confidence interval 1.08 to 2.38). Sensitivity analysis considering the existence of depressive symptoms showed a similar association (hazard ratio 1.62, 1.08 to 2.43). A secondary analysis pooled cohorts of participants who started benzodiazepines during follow-up and evaluated the association with incident dementia. The pooled hazard ratio across the five cohorts of new benzodiazepine users was 1.46 (1.10 to 1.94). Results of a complementary nested case-control study showed that ever use of benzodiazepines was associated with an approximately 50% increase in the risk of dementia (adjusted odds ratio 1.55, 1.24 to 1.95) compared with never users. The results were similar in past users (odds ratio 1.56, 1.23 to 1.98) and recent users (1.48, 0.83 to 2.63) but reached significance only for past users. | | <https://pubmed.ncbi.nlm.nih.gov/23045258/> |
|  |  |  | Fifty patients currently taking benzodiazepines for at least one year, thirty-four who had stopped taking benzodiazepines, and a matched control group of subjects who had never taken benzodiazepines or who had taken benzodiazepines in the past for less than one year were administered a battery of neuropsychological tests designed to measure a wide range of cognitive functions. It was found that patients taking high doses of benzodiazepines for long periods of time perform poorly on tasks involving visual-spatial ability and sustained attention. This is consistent with deficits in posterior cortical cognitive function. | | <https://pubmed.ncbi.nlm.nih.gov/2899898/> |
| **77** | | Antipsychotics are linked to increased stroke and pneumonia risk. | Five hundred ninety patients with ARF (11.7%) filled at least 1 antipsychotic prescription during the case period compared with 443 (8.8%) during the control period, corresponding to a 1.66-fold (95% CI, 1.34-2.05; P < .001) adjusted increased risk of ARF regardless of antipsychotic class and administration route. | | <https://pubmed.ncbi.nlm.nih.gov/28055066/> |
|  |  |  | Monotherapy use was associated with increased pneumonia risk compared with no antipsychotic use (AHR, 1.15 [95% CI, 1.02-1.30]; P = .03) in a dose-dependent manner, but polytherapy use was not. When categorized by anticholinergic burden, only the use of antipsychotics with a high anticholinergic burden was associated with pneumonia (AHR, 1.26 [95% CI, 1.10-1.45]; P < .001). Of specific drugs, high-dose quetiapine (AHR, 1.78 [95% CI, 1.22-2.60]; P = .003), high- and medium-dose clozapine (AHR, 1.44 [95% CI, 1.22-1.71]; P < .001 and AHR, 1.43 [95% CI, 1.18-1.74]; P < .001, respectively), and high-dose olanzapine (AHR, 1.29 [95% CI, 1.05-1.58]; P = .02) were associated with increased pneumonia risk. | | <https://pubmed.ncbi.nlm.nih.gov/38922592/> |
|  |  |  | Compared with non-use, any antipsychotic use was associated with increased risks of all outcomes, except ventricular arrhythmia. Current use (90 days after a prescription) was associated with elevated risks of pneumonia (hazard ratio 2.19, 95% confidence interval (CI) 2.10 to 2.28), acute kidney injury (1.72, 1.61 to 1.84), venous thromboembolism (1.62, 1.46 to 1.80), stroke (1.61, 1.52 to 1.71), fracture (1.43, 1.35 to 1.52), myocardial infarction (1.28, 1.15 to 1.42), and heart failure (1.27, 1.18 to 1.37). No increased risks were observed for the negative control outcome (appendicitis and cholecystitis). In the 90 days after drug initiation, the cumulative incidence of pneumonia among antipsychotic users was 4.48% (4.26% to 4.71%) versus 1.49% (1.45% to 1.53%) in the matched cohort of non-users (difference 2.99%, 95% CI 2.77% to 3.22%). | | <https://pubmed.ncbi.nlm.nih.gov/38631737/> |
| **78** | | Rivastigmine improves cognition/function in AD and PDD. | Cognitive effects were significant for all drugs, ranging from a -1.29 points mean difference (95% CI -2.30 to -0.28) in the 20 mg daily memantine trials to -3.20 points (95% CI -3.28 to -3.12) in the 32 mg daily galantamine group. Only memantine had no effect on the Clinicians' Global Impression of Change scale. No behavioral benefits were observed, except for -2.72 (95% CI -4.92 to -0.52) in the 10 mg daily donepezil group and -1.72 (95% CI -3.12 to -0.33) for 24 mg daily galantamine trial. Only 5 mg daily donepezil had no effect on the function outcome. Compared with placebo, more dropouts and adverse events occurred with the cholinesterase inhibitors, but not with memantine. | | <https://pubmed.ncbi.nlm.nih.gov/24662102/> |
|  |  |  | A total of 541 patients were enrolled, and 410 completed the study. The outcomes were better among patients treated with rivastigmine than among those who received placebo; however, the differences between these two groups were moderate and similar to those reported in trials of rivastigmine for Alzheimer's disease. Rivastigmine-treated patients had a mean improvement of 2.1 points in the score for the 70-point ADAS-cog, from a baseline score of 23.8, as compared with a 0.7-point worsening in the placebo group, from a baseline score of 24.3 (P<0.001). | | <https://pubmed.ncbi.nlm.nih.gov/15590953/> |
|  |  |  | At 48 weeks, the mean ADAS-cog score for the whole group improved by 2 points above baseline. Placebo patients switching to rivastigmine for the active treatment extension experienced a mean cognitive improvement similar to that of the original rivastigmine group during the double-blind trial. The adverse event profile was comparable to that seen in the double-blind trial. Long-term rivastigmine treatment appeared well tolerated and may provide sustained benefits in dementia associated with PD patients who remain on treatment for up to 48 weeks. | | <https://pubmed.ncbi.nlm.nih.gov/16229010/> |
| **79** | | Default mode network connectivity is disrupted in MCI and AD. | Thirty-four studies were included (1363 participants, average 40 per study). Consistent alterations in connectivity were found in the default mode, salience, and limbic networks in patients with AD dementia, mild cognitive impairment, or in both groups. We also identified a strong tendency in the literature toward specific examination of the default mode network. | | <https://pubmed.ncbi.nlm.nih.gov/28560308/> |
|  |  |  | Qualitatively, our review of 57 MCI versus HC comparisons suggests substantial inconsistency; 9 showed no group difference, 8 showed MCI > HC and 22 showed HC > MCI across the brain, and 18 showed regionally-mixed directions of effect. The meta-analysis of 31 studies revealed areas of significant hypo- and hyper-connectivity in MCI, including hypoconnectivity in the posterior cingulate cortex/precuneus (z = -3.1, p < 0.0001). Very few individual studies, however, showed patterns resembling the meta-analytic results. Methodological differences did not appear to explain inconsistencies. | | <https://pubmed.ncbi.nlm.nih.gov/31177210/> |
|  |  |  | Within-DMN connectivity is a marker of episodic memory performance even among cognitively healthy older adults. | | <https://pubmed.ncbi.nlm.nih.gov/29440553/> |
| **80** | | Unlike midlife obesity, rapid weight loss in older adults indicates progression of cognitive decline. | Compared to participants with stable weight, those with weight loss had increased odds of diagnostic progression (adjusted OR= 1.35, 95%CI [1.21, 1.51]). Also, large weight fluctuation was associated with increased odds of diagnostic progression (OR comparing the extreme quartiles = 1.20, 95%CI [1.04, 1.39]) after adjusting for traditional risk factors for dementia and body weight change. The magnitude of the association appeared larger among those older than 80 and those with 3 or more cardiometabolic risk factors at baseline (both p for interaction <0.05). | | <https://pmc.ncbi.nlm.nih.gov/articles/PMC8759073/> |
|  |  |  | Higher late-life BMI is associated with a lower risk of incident MCI and AD but is not protective in the presence of rapid weight loss. | | <https://pmc.ncbi.nlm.nih.gov/articles/PMC5736008/> |
|  |  |  | BMI loss was associated with an increased risk of all-cause dementia and Alzheimer's disease in participants without obesity; however, this association was absent in participants with obesity. | | <https://pubmed.ncbi.nlm.nih.gov/37216633/> |
| **81** | | Olfactory impairment is asssociated with incident dementia and faster progression from MCI to AD. | In logistic regression analyses, lower baseline UPSIT scores were associated with cognitive decline (relative risk 1.067 per point interval; 95% confidence interval [CI] 1.040, 1.095; p < 0.0001), and remained significant (relative risk 1.065 per point interval; 95% CI 1.034, 1.095; p < 0.0001) after including covariates. UPSIT, but not Selective Reminding Test-total immediate recall, predicted cognitive decline in participants without baseline cognitive impairment. During follow-up, 101 participants transitioned to AD dementia. In discrete time survival analyses, lower baseline UPSIT scores were associated with transition to AD dementia (hazard ratio 1.099 per point interval; 95% CI 1.067, 1.131; p < 0.0001), and remained highly significant (hazard ratio 1.072 per point interval; 95% CI 1.036, 1.109; p < 0.0001) after including demographic, cognitive, and functional covariates. | | <https://pubmed.ncbi.nlm.nih.gov/25471394/> |
|  |  |  | In a proportional hazards model adjusted for age, sex, and education, odor identification score predicted development of MCI (relative risk, 1.15; 95% confidence interval, 1.07-1.23), with risk increased by 50% in persons with below-average (score of 8 [25th percentile]) compared with above-average (score of 11 [75th percentile]) odor identification scores. Results were not substantially changed in subsequent analyses that controlled for level of cognitive function or disability, presence of stroke, or smoking status at baseline or that required MCI to persist for at least 1 year. Impaired odor identification was also associated with a lower level of global cognition at baseline and with more rapid decline in episodic memory, semantic memory, and perceptual speed. | | <https://pubmed.ncbi.nlm.nih.gov/17606814/> |
|  |  |  | Faster olfactory decline during periods of normal cognition predicted higher incidence of subsequent MCI or dementia (OR 1.89, 95% CI: 1.26, 2.90, p < 0.01; comparable to carrying an APOE-ε4 allele) and smaller GMV in AD and olfactory regions (β = -0.11, 95% CI -0.21, -0.00). | | <https://pubmed.ncbi.nlm.nih.gov/35899859/> |
| **82** | | Mild behavioral impairment (MBI) is predicts conversion to AD or other dementias. | MBI and SCD together were associated with the greatest risk of decline. These complementary dementia risk syndromes can be used as simple and scalable methods to identify high-risk patients for workup or for clinical trial enrichment. | | <https://pubmed.ncbi.nlm.nih.gov/33554909/> |
|  |  |  | Mild behavioral impairment (MBI) can identify older adults at risk of dementia. Neuropsychiatric symptom (NPS) assessment tools can be proxy measures for MBI.Hazard for dementia was highest for MBI defined by NPS presence at more than two-thirds of visits. | | <https://pubmed.ncbi.nlm.nih.gov/37786862/> |
|  |  |  | MCI patients mostly converted to Alzheimer's dementia, while MBI converted to frontotemporal dementia and Lewy body dementia. Patients in PG converted to Lewy body dementia and frontotemporal dementia. | | <https://pubmed.ncbi.nlm.nih.gov/29439333/> |
| **83** | | History of traumatic brain injury is associated with earlier onset and higher risk of dementia. | The study included 178 779 patients diagnosed with a TBI in the Veterans Health Administration health care system and 178 779 patients in a propensity-matched comparison group. Veterans had a mean (SD) age of nearly 49.5 (18.2) years at baseline; 33 250 (9.3%) were women, and 259 136 (72.5%) were non-Hispanic white individuals. Differences between veterans with and without TBI were small. A total of 4698 veterans (2.6%) without TBI developed dementia compared with 10 835 (6.1%) of those with TBI. After adjustment for demographics and medical and psychiatric comobidities, adjusted hazard ratios for dementia were 2.36 (95% CI, 2.10-2.66) for mild TBI without LOC, 2.51 (95% CI, 2.29-2.76) for mild TBI with LOC, 3.19 (95% CI, 3.05-3.33) for mild TBI with LOC status unknown, and 3.77 (95% CI, 3.63-3.91) for moderate to severe TBI. | | <https://pubmed.ncbi.nlm.nih.gov/29801145/> |
|  |  |  | Among patients evaluated in the ED or inpatient settings, those with moderate to severe TBI at 55 years or older or mild TBI at 65 years or older had an increased risk of developing dementia. Younger adults may be more resilient to the effects of recent mild TBI than older adults. | | <https://pubmed.ncbi.nlm.nih.gov/25347255/> |
|  |  |  | During the 5 year follow-up period, 1196 TBI (2.66%) and 224,625 non-TBI patients (1.53%) patients developed dementia. During the 5 year follow-up period, TBI was independently associated with a 1.68 (range 1.57-1.80) times greater risk of dementia after adjusting for sociodemographic characteristics and selected comorbidities. | | <https://pubmed.ncbi.nlm.nih.gov/22842203/> |
| **84** | | Higher frailty burden predicts incident dementia and faster decline. | Frailty was a significant predictor of incident Alzheimer disease (4 studies: pooled HR = 1.28, 95% confidence interval (95% CI) = 1.00-1.63, P = .05), vascular dementia (2 studies: pooled HR 2.70, 95% CI 1.40-5.23, P = .003), and all dementia (3 studies: pooled HR 1.33, 95% CI 1.07-1.67, P = .01). Heterogeneity across the studies was low to modest (I(2) = 0%-51%). A random-effects meta-regression analysis showed that the female proportion of the cohort primarily mediated the association of frailty with Alzheimer disease (female proportion coefficient = 0.04, 95%CI = 0.01-0.08, P = .01). | | <https://pubmed.ncbi.nlm.nih.gov/27324809/> |
|  |  |  | Frailty was associated with cumulative incidence of dementia (p<0.001), MMSE score at baseline (p<0.001), difference in MMSE score between 2009 and 2014 (p 0.005), CIRS (p<0.001).After four follow-up years, 4.6% developed Dementia. In the longitudinal regression models adjusted for age, gender, level of education and loneliness, APOEɛ4 carrier status, Frailty was associated with incident dementia (OR=7.75,95% CI 3.26-18.41), with higher risk of dementia for frail men compared to women (OR=13.79, 95% CI 4.11–46.21; OR= 4.63, 95% CI 1.32-16.26, respectively). | | <https://alz-journals.onlinelibrary.wiley.com/doi/10.1002/alz.042509> |
|  |  |  | Among 1931 participants, 348 (18.0%) were capable of sustained frailty remission. During the 8-year follow-up, 279 participants developed dementia. In a fully adjusted model, sustained remission was associated with a lower risk of dementia (hazard ratio = 0.66, 95% confidence interval = 0.47 to 0.93). The association was more pronounced among younger-old and male participants but not observed among their counterparts. | | <https://pubmed.ncbi.nlm.nih.gov/39016447/> |
| **85** | | Hippocampal atrophy is more pronounced in Alzheimer’s disease than in Parkinson’s disease dementia. | In addition, significant temporal lobe atrophy, including the hippocampus and parahippocampal gyrus was detected in Alzheimer's disease relative to PDD. | | <https://pubmed.ncbi.nlm.nih.gov/14749292/> |
|  |  |  | Medial temporal lobe atrophy (MTA) was seen in cognitively intact older subjects with Parkinson disease (PD) and was not more pronounced in Parkinson disease dementia (PDD). Alzheimer disease (AD) and, to a lesser extent, dementia with Lewy bodies (DLB) showed more pronounced MTA. | | <https://pubmed.ncbi.nlm.nih.gov/15753423/> |
|  |  |  | AD showed atrophy and increased p-tau, but not amyloid-β, burden in the CA1, subiculum and entorhinal cortex compared to controls, however MRI and neuropathology did not correlate. Controls and PD had similar hippocampal subfield volumes and pathology load. | | <https://pubmed.ncbi.nlm.nih.gov/40448161/> |
| **86** | | Delirium is a predictor of subsequent cognitive decline across clinical populations. | In all studies, the group that experienced delirium had worse cognition at the final time point. The I2 measure of between-study variability in g was 0.81. A multivariable meta-regression suggested that duration of follow-up (longer with larger gs), number of covariates controlled (greater numbers were associated with smaller gs), and baseline cognitive matching (matching was associated with larger gs) were significant sources of variance. More specialized subgroup and meta-regressions were consistent with predictions that suggested that delirium may be a causative factor in cognitive decline. | | <https://pubmed.ncbi.nlm.nih.gov/32658246/> |
|  |  |  | With hospitalization and delirium, the adjusted RR for cognitive decline for patients with AD was 1.6 (CI, 1.2 to 2.3). Among hospitalized patients with AD, 21% of the incidences of cognitive decline, 15% of institutionalization, and 6% of deaths were associated with delirium. | | <https://pubmed.ncbi.nlm.nih.gov/22711077/> |
|  |  |  | During the 2-year follow-up, 89 (42%) of 210 participants were diagnosed with one or more episodes of delirium. Incident delirium was independently associated with a decrease in MoCA score at the next 6-month follow-up, even after adjustment for age, sex, education, previous MoCA score, and frailty (adjusted mean difference –1·5, 95% CI –2·9 to –0·1). We found an interaction between previous MoCA score and delirium (β –0·254, 95% CI –0·441 to –0·066, p=0·010), with the largest decline being observed in people with better baseline cognition. Participants with delirium and good previous cognitive function and participants with a high peak concentration of NfL during any hospitalisation had increased NfL at the next 6-month follow-up. Mediation analyses showed independent pathways from previous MoCA score to follow-up MoCA score with contributions from incident delirium (–1·7, 95% CI –2·8 to –0·6) and from previous NfL to follow-up MoCA score with contributions from acute NfL concentrations (–1·8, –2·5 to –1·1). | | <https://pubmed.ncbi.nlm.nih.gov/37459878/> |
| **87** | | Vitamin B12 deficiency is associated with brain atrophy. | In the hippocampal atrophy group, the frequency of vitamin B12 deficiency was higher (p < 0.022), MMSE score was lower (p < 0.0001), and age was higher (p < 0.0001) than that in the normal group (Mann-Whitney U test). Patients with vitamin B12 deficiency (odds ratio, 3.46) and low MMSE score (odds ratio, 2.24) had an increased risk of hippocampal atrophy. | | <https://pubmed.ncbi.nlm.nih.gov/40675920/> |
|  |  |  | Using the upper (for the vitamins) or lower tertile (for the metabolites) as reference in logistic regression analysis and adjusting for the above covariates, vitamin B(12) in the bottom tertile (<308 pmol/L) was associated with increased rate of brain volume loss (odds ratio 6.17, 95% CI 1.25-30.47). | | <https://pubmed.ncbi.nlm.nih.gov/18779510/> |
|  |  |  | Concentrations of all vitamin B12-related markers, but not serum vitamin B12 itself, were associated with global cognitive function and with total brain volume. Methylmalonate levels were associated with poorer episodic memory and perceptual speed, and cystathionine and 2-methylcitrate with poorer episodic and semantic memory. Homocysteine concentrations were associated with decreased total brain volume. | | <https://pubmed.ncbi.nlm.nih.gov/21947532/> |
| **88** | | Heart failure is associated with an increased risk of dementia in older adults. | During the 6534 person-years of follow-up (mean, 5.02 years per person), 440 subjects were diagnosed as having dementia, including 333 with Alzheimer disease. At baseline, heart failure was identified in 205 subjects. Heart failure was associated with a multi-adjusted hazard ratio (HR) of 1.84 (95% confidence interval [CI], 1.35-2.51) for dementia and 1.80 (95% CI, 1.25-2.61) for Alzheimer disease. | | <https://pubmed.ncbi.nlm.nih.gov/16682574/> |
|  |  |  | We identified 16 studies (1,309,483 individuals) regarding CHD, and seven studies (1,958,702 individuals) about HF. A history of CHD was associated with a 27% increased risk of dementia (pooled relative risk [RR] [95% confidence interval, CI]: 1.27 [1.07–1.50]), albeit with considerable heterogeneity across studies (I2 = 80%). HF was associated with 60% increased dementia risk (pooled RR 1.60 [1.19–2.13]) with moderate heterogeneity (I2 = 59%). Among prospective population-based cohorts, pooled estimates were similar (for CHD, RR 1.26 [1.06–1.49], nine studies; and HF, RR 1.80 [1.41–2.31], four studies) and highly consistent (I2 = 0%). | | <https://pubmed.ncbi.nlm.nih.gov/29494808/> |
|  |  |  | AF in late-life was an independent risk factor for dementia (HR 2.61, 95% CI 1.05–6.47; p = 0.039) and AD (HR 2.54, 95% CI 1.04–6.16; p = 0.040) in the fully adjusted analyses. The association was even stronger among the apolipoprotein E (APOE) ε4 non-carriers. Late-life HF, but not CAD, tended to increase the risks as well. Heart diseases diagnosed at midlife did not increase the risk of later dementia and AD. | | <https://pubmed.ncbi.nlm.nih.gov/24825565/> |
| **89** | | Having epilepsy is associated with increased risk of dementia. | Participants had a mean (SD) age of 54.3 (5.8) years at baseline, 57.7% were female, 28.2% were of self-reported Black race, 14.4% were ultimately categorized as having head injury, 5.1% as having seizure/epilepsy, and 1.2% as having PTE. Over a median follow-up of 25 (25th to 75th percentile, 17-30) years, 19.9% developed dementia. In fully adjusted models, compared with no head injury and no seizure/epilepsy, PTE was associated with 4.56 (95% CI, 4.49-5.95) times the risk of dementia, while seizure/epilepsy was associated with 2.61 (95% CI, 2.21-3.07) times the risk and head injury with 1.63 (95% CI, 1.47-1.80) times the risk. The risk of dementia associated with PTE was significantly higher than the risk associated with head injury alone and with nontraumatic seizure/epilepsy alone. | | <https://pubmed.ncbi.nlm.nih.gov/38407883/> |
|  |  |  | Of 495 149 participants (225 481 [45.5%] men; mean [SD] age, 57.5 [8.1] years), 3864 had a diagnosis of focal epilepsy only, 6397 had a history of stroke only, and 14 518 had migraine only. Executive function was comparable between participants with epilepsy and stroke and worse than the control and migraine group. Focal epilepsy was associated with a higher risk of developing dementia (hazard ratio [HR], 4.02; 95% CI, 3.45 to 4.68; P < .001), compared with stroke (HR, 2.56; 95% CI, 2.28 to 2.87; P < .001), or migraine (HR, 1.02; 95% CI, 0.85 to 1.21; P = .94). Participants with focal epilepsy and high cardiovascular risk were more than 13 times more likely to develop dementia (HR, 13.66; 95% CI, 10.61 to 17.60; P < .001) compared with controls with low cardiovascular risk. The imaging subsample included 42 353 participants. Focal epilepsy was associated with lower hippocampal volume (mean difference, −0.17; 95% CI, −0.02 to −0.32; t = −2.18; P = .03) and lower total gray matter volume (mean difference, −0.33; 95% CI, −0.18 to −0.48; t = −4.29; P < .001) compared with controls. There was no significant difference in white matter hyperintensity volume (mean difference, 0.10; 95% CI, −0.07 to 0.26; t = 1.14; P = .26). | | <https://pubmed.ncbi.nlm.nih.gov/36972059/> |
|  |  |  | Of 9,033 ARIC participants with sufficient Medicare coverage data (4,980 [55.1%] female, 1993 [22.1%] Black), 671 met the definition of LOE. Two hundred seventy-nine (41.6%) participants with and 1,408 (16.8%) without LOE developed dementia (p < 0.001). After a diagnosis of LOE, the adjusted hazard ratio for developing subsequent dementia was 3.05 (95% confidence interval 2.65-3.51). The median time to dementia ascertainment after the onset of LOE was 3.66 years (quartile 1-3, 1.28-8.28 years). | | <https://pubmed.ncbi.nlm.nih.gov/33097597/> |
| **90** | | Greater EEG slowing is associated with higher risk of conversion to dementia. | Associations between qEEG slowing, measured by increased theta/alpha ratio, and clinical progression from MCI to dementia were estimated with a multistate transition model to account for death as a competing risk, while controlling for age, cognitive function, and etiology classified by an expert consensus panel.Over a mean follow-up of 1.5 years (SD = 0.5), 14 cases of incident dementia and 5 deaths were observed. Increased theta/alpha ratio on qEEG was associated with increased annual hazard of dementia (hazard ratio = 1.84, 95% CI: 1.01-3.35). This extends previous findings that MCI-LB features early functional changes, showing that qEEG slowing may anticipate the onset of dementia in prospectively identified MCI. | | <https://pubmed.ncbi.nlm.nih.gov/34551831/> |
|  |  |  | In the higher alpha3/alpha2 frequency power ratio group, greater memory impairment was correlated with greater cortical atrophy and lower perfusional rate in the temporo-parietal cortex. After a follow-up of three years, these patients converted in AD. | | <https://pubmed.ncbi.nlm.nih.gov/26715588/> |
|  |  |  | Compared with participants in the lowest quartile of TAR, those in the highest quartile had a 2.2-fold risk of developing dementia in MrOS and a 10.0-fold risk of developing dementia in SOF (P’s <0·01). Furthermore, a higher TAR at baseline and greater increase in TAR over time, were associated with greater genetic risk for dementia, faster cognitive decline, faster epigenetic ageing measured by DunedinPACE, and increased risk of all-cause mortality (P’s <0·05). | | <https://www.neurology.org/doi/10.1212/WNL.0000000000208935> |
| **91** | | Bipolar disorder is associated with increased risk of dementia. | This study included 501,233 UK Biobank participants (mean [standard deviation] age, 56.5 [8.10] years; 54.4% women), free of dementia and PD at baseline. After a median 13.8 year follow-up, 9422 cases of dementia and 3457 PD cases were identified. Participants with BD had a significantly higher risk of dementia (adjusted hazard ratio [HR] 2.52, 95% CI 2.00–3.19) and PD (adjusted HR 2.88, 95% CI 2.03-4.08). | | <https://pubmed.ncbi.nlm.nih.gov/39362870/> |
|  |  |  | Bipolar disorder was associated with increased adjusted hazard ratio (HR) of dementia (HR = 2.30, 95% CI 1.80–2.94). The risk of dementia was greatest among those with <5 years of history of bipolar disorder or who had had illness onset after 70 years of age. Bipolar disorder was also associated with increased mortality (HR = 1.51, 95% CI 1.28–1.77). | | <https://pubmed.ncbi.nlm.nih.gov/27482038/> |
|  |  |  | Patients with unipolar or bipolar affective disorder seem to have an increased risk of developing dementia compared to patients with other illnesses. | | <https://pubmed.ncbi.nlm.nih.gov/12547295/> |
| **92** | | Schizophrenia is associated with increased risk of dementia. | During 18 years of follow-up, 136 012 individuals, including 944 individuals with a history of schizophrenia, developed dementia. Schizophrenia was associated with a more than 2-fold higher risk of all-cause dementia (IRR, 2.13; 95% CI, 2.00-2.27) after adjusting for age, sex, and calendar period. The estimates (reported as IRR; 95% CI) did not change substantially when adjusting for medical comorbidities, such as cardiovascular diseases and diabetes mellitus (2.01; 1.89-2.15) but decreased slightly when adjusting for substance abuse (1.71; 1.60-1.82). The association between schizophrenia and dementia risk was stable when evaluated in subgroups characterized by demographics and comorbidities, although the IRR was higher among individuals younger than 65 years (3.77; 3.29-4.33), men (2.38; 2.13-2.66), individuals living with a partner (3.16; 2.71-3.69), those without cerebrovascular disease (2.23; 2.08-2.39), and those without substance abuse (1.96; 1.82-2.11). The CIPs (95% CIs) of developing dementia by the age of 65 years were 1.8% (1.5%-2.2%) for persons with schizophrenia and 0.6% (0.6%-0.7%) for persons without schizophrenia. | | <https://pubmed.ncbi.nlm.nih.gov/26444987/> |
|  |  |  | The study population of 8 011 773 adults 66 years or older (63.4% women; mean [SD] age, 74.0 [8.2] years) included 74 170 individuals with a diagnosis of schizophrenia (56.6% women) and 7 937 603 without an SMI diagnosis (63.5% women) who contributed 336 814 and 55 499 543 person-years of follow-up, respectively. At 66 years of age, the prevalence of diagnosed dementia was 27.9% (17 640 of 63 287) among individuals with schizophrenia compared with 1.3% (31 295 of 2 389 512) in the group without SMI. By 80 years of age, the prevalence of dementia diagnoses was 70.2% (2011 of 2866) in the group with schizophrenia and 11.3% (242 094 of 2 134 602) in the group without SMI. The annual incidence of dementia diagnoses per 1000 person-years at 66 years of age was 52.5 (95% CI, 50.1-54.9) among individuals with schizophrenia and 4.5 (95% CI, 4.4-4.6) among individuals without SMI and increased to 216.2 (95% CI, 179.9-252.6) and 32.3 (95% CI, 32.0-32.6), respectively, by 80 years of age. | | <https://pubmed.ncbi.nlm.nih.gov/33688938/> |
|  |  |  | We found a substantially higher rate of dementia in individuals with VLOSLP [hazard ratio (HR): 4.22, 95% confidence interval (95% CI) 4.05–4.41]. Median time-to-dementia-diagnosis was 75% shorter in those with VLOSLP (time ratio: 0.25, 95% CI 0.24–0.26). This association was strongest in the first year following VLOSLP diagnosis, and attenuated over time, although dementia rates remained higher in participants with VLOSLP for up to 20 years of follow-up. | | <https://pubmed.ncbi.nlm.nih.gov/34030750/> |
| **93** | | Thyroid disease is associated with higher risk of dementia. | Among patients aged 65 years or older, a history of hypothyroidism was associated with an increased risk of being diagnosed with dementia (adjusted odds ratio [aOR] 1.81; 95% CI 1.14-2.87; p = 0.011), which was an association not present in patients older than 50 years but younger than 65 years. We found that this association was most significant among patients aged 65 years or older with a history of hypothyroidism who received hypothyroidism medication (aOR 3.17; 95% CI 1.04-9.69; p = 0.043). | | <https://pubmed.ncbi.nlm.nih.gov/35794019/> |
|  |  |  | During a mean follow-up of 12.7 years (range, 1-25 years), 209 participants (142 women) developed AD. Women in the lowest (<1.0 mIU/L) and highest (>2.1 mIU/L) tertiles of serum thyrotropin concentration were at increased risk for AD (multivariate-adjusted hazard ratio, 2.39 [95% confidence interval, 1.47-3.87] [P < .001] and 2.15 [95% confidence interval, 1.31-3.52] [P = .003], respectively) compared with those in the middle tertile. In analyses limited to participants with serum thyrotropin levels of 0.1 to 10.0 mIU/L, the U-shaped relationship between thyrotropin level and AD risk was maintained in women but not when analyses were limited to those with thyrotropin levels of 0.5 to 5.0 mIU/L. | | <https://pubmed.ncbi.nlm.nih.gov/18663163/> |
|  |  |  | . Higher total and free thyroxine levels were associated with an increased risk of dementia and AD (age and sex adjusted hazard ratio (95% confidence interval) per S.D. increase in free thyroxine: 1.21 (1.04; 1.40) and 1.31 (1.14; 1.51), respectively). | | <https://pubmed.ncbi.nlm.nih.gov/17870208/> |
| **94** | | Autism spectrum disorder is associated with higher risk of dementia. | The 5-Year prevalence of dementia was 4.04% among adults with ASD only, and 5.22% for those with ASD and co-occurring ID. This prevalence was higher compared to the prevalence of dementia in individuals with no ASD and no ID (0.97%), but lower compared to individuals with ID only (7.10%). Risk factors associated with the increased prevalence in the general population were similarly associated with the increased risk of dementia in individuals with ASD. Even after adjusting for these risk factors, compared to the general population, dementia was found to occur more frequently in individuals with ASD only (adjusted hazard ratio, 1.96; 95% CI, 1.69–2.28), as well as individuals with ASD and co-occurring ID (adjusted hazard ratio, 2.89; 95% CI, 2.62–3.17). | | <https://pmc.ncbi.nlm.nih.gov/articles/PMC8487995/> |
|  |  |  | Linked Medicaid and Medicare records suggest a markedly elevated prevalence of identified dementia diagnoses in individuals with an ASD diagnosis. Findings were consistent with our previous research, and prevalence estimates were higher than those for the general population of Medicaid and Medicare beneficiaries reported in the literature. | | <https://pmc.ncbi.nlm.nih.gov/articles/PMC11696448/> |
|  |  |  | "We calculated prevalence, incidence, age at onset, and created survival curves. There were 90,229 autistic adults ≥ 30 years of age and enrolled for at least 1 year in Medicaid and/or Medicare and 267 ADRD cases. Prevalence of ADRD was 2.09% (95% CI: 1.99%, 2.20%) in 2011 and 8.11% (95% CI: 7.92%, 8.30%) in 2019. Mean age at ADRD onset was 59.3 years (SD: 14.2). Mean age among men was 58.3 years (SD: 13.8) and 61.0 years among females. Incidence of ADRD was higher in autistic adults with intellectual disability with no difference by sex. ADRD is a prevalent condition in middle- and older-aged adults identified with autism in the Medicaid and Medicare system." "Prevalence and incidence of ADRD was higher in the autistic population compared to studies of the general population." | | <https://pubmed.ncbi.nlm.nih.gov/40166852/> |
| **95** | | Higher cardiorespiratory fitness is associated with lower dementia risk. | Participants who had an increased estimated cardiorespiratory fitness over time had a reduced risk of incident dementia (AHR 0·52, 95% CI 0·30-0·90) and dementia mortality (0·72, 0·52-0·99) when compared with those who remained unfit at both assessments. | | <https://pubmed.ncbi.nlm.nih.gov/31677775/> |
|  |  |  | Compared to the least-fit, multivariable-adjusted hazard ratios (95% confidence intervals) for incident ADRD were: 0.87 (0.85-0.90), 0.80 (0.78-0.83), 0.74 (0.72-0.76), and 0.67 (0.65-0.70), for low-fit, moderate-fit, fit, and high-fit individuals, respectively. | | <https://pubmed.ncbi.nlm.nih.gov/36946469/> |
|  |  |  | Increases in cardiorespiratory fitness were associated with increases in brain activation in both the left inferior frontal and precentral gyri. Furthermore, changes in cardiorespiratory fitness were also correlated with changes in performance on several neuropsychological tests. | | <https://pubmed.ncbi.nlm.nih.gov/33720893/> |
| **96** | | Reduced hippocampal volume correlates with worse memory performance. | Decreased hippocampal volume was strongly associated with worse performance in total recall, and lower entorhinal cortex CBV was associated with lower performance in delayed recall. | | <https://pubmed.ncbi.nlm.nih.gov/19901171/> |
|  |  |  | Multivariate conditional inference analysis showed that gender and left hippocampal volume largely dominated predictive values for CVLT-LD scores in our sample. Left hippocampal volume dominated predictive values for females but not for males. | | <https://pubmed.ncbi.nlm.nih.gov/19698138/> |
|  |  |  | The volume of the right hippocampus (r = 0.37, n = 32, p <0.05) and the magnitude of the asymmetry between the right and left hippocampi (r = 0.38, n = 32, p <0.05) correlated with total score on the Benton test. We also found significant correlations between the amygdaloid volumes and the performance on visual memory tests but not with score on the verbal memory test. | | <https://www.neurology.org/doi/abs/10.1212/wnl.44.9.1660?casa_token=vOdxtR4QBCoAAAAA:UsF9eofmLRuiq_biSh6uoGGBPtRAu-6VM0f4Tx8y9DqfAjV33CvEoUJyZMHOrfj3LnYscLyS15hu> |
| **97** | | Maternal history of dementia is more associated with the risk of dementia compared to paternal history of dementia. | In this cross-sectional study of 4413 individuals screened for the Anti-Amyloid Treatment in Asymptomatic Alzheimer (A4) study, individuals with a maternal history of memory impairment had elevated neocortical β-amyloid cross-sectionally compared with individuals with only paternal history or no parental history. | | <https://pmc.ncbi.nlm.nih.gov/articles/PMC11184498/> |
|  |  |  | A family history of Alzheimer's disease (AD) increases one's risk of developing late-onset AD (LOAD), and a maternal family history of LOAD influences risk more than a paternal family history. Accumulating evidence suggests that a family history of dementia associates with AD-typical biomarker changes. We analyzed cross-sectional data from non-demented, mild cognitive impairment (MCI), and LOAD participants in the Alzheimer's Disease Neuroimaging Initiative (ADNI) with PET imaging using Pittsburgh Compound B (PiB, n = 99) and cerebrospinal fluid (CSF) analysis (n = 403) for amyloid-β peptide (Aβ) and total tau. We assessed the relationship of CSF and PiB biomarkers and family history of dementia, as well as parent gender effects. In the larger analysis of CSF biomarkers, we assessed diagnosis groups individually. In the overall sample, CSF Aβ, tau/Aβ ratio, and global PiB uptake were significantly different between family history positive and negative groups, with markers of increased AD burden associated with a positive maternal family history of dementia. Moreover, a maternal family history of dementia was associated with significantly greater PiB Aβ load in the brain in the parietal cortex, precuneus, and sensorimotor cortex. | | <https://pubmed.ncbi.nlm.nih.gov/22669011/> |
|  |  |  | The mean age of the participants was 72.8 ± 7.9 years and 59.2% were female. Parental history of dementia was associated with higher risk of dementia (odds ratio [OR] = 1.47, 95% confidence interval [CI] = 1.15–1.86) and Alzheimer's disease (AD) (OR = 1.72, 95% CI = 1.31–2.26), but not with the risk of non-AD. This was largely driven by maternal history of dementia, which was associated with the risk of dementia (OR = 1.51, 95% CI = 1.15–1.97) and AD (OR = 1.80, 95% CI = 1.33–2.43) whereas paternal history of dementia was not. These results remained significant when males and females were analyzed separately (OR = 2.14, 95% CI = 1.28–3.55 in males; OR = 1.68, 95% CI = 1.16–2.44 for females). | | <https://pubmed.ncbi.nlm.nih.gov/37165609/> |
| **98** | | Depression is associated with lower hippocampal volume. | : Hippocampal volume is reduced in patients with unipolar depression, maybe as a consequence of repeated periods of major depressive disorder. | | <https://pubmed.ncbi.nlm.nih.gov/15514393/> |
|  |  |  | Smaller hippocampal volumes were strongly associated with poorer verbal learning and memory as well as diagnoses of either multiple or amnestic mild cognitive impairment. Based on univariate correlations, multivariable regressions were performed (controlling for age and total intracranial volume) to determine which modifiable risk factors were associated with hippocampal volume. For the left hippocampus, poor sleep efficiency and greater than five years untreated depressive illness remained significant predictors. | | <https://pubmed.ncbi.nlm.nih.gov/25408219/> |
|  |  |  | Longer durations during which depressive episodes went untreated with antidepressant medication were associated with reductions in hippocampal volume. | | <https://pubmed.ncbi.nlm.nih.gov/12900317/> |
| **99** | | Air pollution exposure is associated with increased dementia risk. | In meta-analyses of incident dementia, we identified a dementia diagnosis to be significantly associated with long-term exposure to PM2·5 (21 studies, n=24 030 527, pooled adjusted hazard ratio (HR) per 5 μg/m3 increase in exposure, 1·08 [95% CI 1·02–1·14]; I2=95%), nitrogen dioxide (16 studies, n=17 228 429, pooled adjusted HR per 10 μg/m3 increase, 1·03 [1·01–1·05]; I2=84%), and black carbon/PM2·5 absorbance (six studies, n=19 421 865, pooled adjusted HR per 1 μg/m3 increase, 1·13 [1·01–1·27]; I2=97%). We found no significant association for exposure to nitrogen oxides (five studies, n=241 409, pooled adjusted HR per 10 μg/m3 increase, 1·05 [0·97–1·13]; I2=44%), PM10 (four studies, n=246 440, pooled adjusted HR per 15 μg/m3 increase, 1·52 [0·80–2·87]; I2=82%), or annual ozone (four studies, n=419 972, pooled adjusted HR per 45 μg/m3 increase, 0·82 [0·35–1·92]; I2=69%), with moderate to considerable heterogeneity between studies in these pooled analyses. | | <https://pubmed.ncbi.nlm.nih.gov/40716448/> |
|  |  |  | Here we report a meta-analysis of 28 longitudinal cohort studies published up to June 2023 that investigated long-term PM2.5 exposure and dementia outcomes. We derived risk–outcome scores (ROSs), highly conservative measures of effect size and evidence strength, mapped onto a 1–5-star rating from ‘weak and/or inconsistent evidence’ to ‘very strong and/or consistent evidence’. We identified a significant nonlinear relationship between PM2.5 exposure and dementia, with a minimum 14% increased risk averaged across PM2.5 levels between 4.5 and 26.9 µg m−3 (the 15th to 85th percentile exposure range across included studies), relative to a reference of 2.0 µg m−3 (n = 49, ROS = 0.13, two stars). We found a significant association of PM2.5 with Alzheimer’s disease (n = 12, ROS = 0.32, three stars) but not with vascular dementia. Our findings highlight the potential impact of air pollution on brain aging. | | <https://pubmed.ncbi.nlm.nih.gov/40119171/> |
|  | |  | 2080 records identified 51 studies for inclusion. Most studies were at high risk of bias, although in many cases bias was towards the null. 14 studies could be meta-analysed for particulate matter <2.5 µm in diameter (PM2.5). The overall hazard ratio per 2 μg/m3 PM2.5 was 1.04 (95% confidence interval 0.99 to 1.09). The hazard ratio among seven studies that used active case ascertainment was 1.42 (1.00 to 2.02) and among seven studies that used passive case ascertainment was 1.03 (0.98 to 1.07). The overall hazard ratio per 10 μg/m3 nitrogen dioxide was 1.02 ((0.98 to 1.06); nine studies) and per 10 μg/m3 nitrogen oxide was 1.05 ((0.98 to 1.13); five studies). Ozone had no clear association with dementia (hazard ratio per 5 μg/m3 was 1.00 (0.98 to 1.05); four studies). | | <https://pubmed.ncbi.nlm.nih.gov/37019461/> |
| **100** | | Greater impairment on the Wisconsin Card Sorting Test is associated with reduced activation or perfusion in frontal-executive cortical regions. | These findings augment evidence that the rostrodorsal prefrontal cortex crucially mediates attentional set shifting, and suggest that the stuck-in-set perseverative errors would be a true pathognomonic sign of frontal dysfunction. Moreover, this study shows that the recurrent perseverative errors may not be associated closely with the prefrontal function, suggesting that this error and the stuck-in-set error should be differentially estimated in the WCST. | | <https://pubmed.ncbi.nlm.nih.gov/15654026/> |
|  |  |  | The results demonstrated specific involvement of different prefrontal areas during different stages of task performance. The mid-dorsolateral prefrontal cortex (area 9/46) increased activity while subjects received either positive or negative feedback, that is at the point when the current information must be related to earlier events stored in working memory. This is consistent with the proposed role of the mid-dorsolateral prefrontal cortex in the monitoring of events in working memory. By contrast, a cortical basal ganglia loop involving the mid-ventrolateral prefrontal cortex (area 47/12), caudate nucleus, and mediodorsal thalamus increased activity specifically during the reception of negative feedback, which signals the need for a mental shift to a new response set. The posterior prefrontal cortex response was less specific; increases in activity occurred during both the reception of feedback and the response period, indicating a role in the association of specific actions to stimuli. | | <https://pubmed.ncbi.nlm.nih.gov/11567063/> |
|  |  |  | Hypoperfusions of the bilateral posterior cingulate, rostrodorsal prefrontal, and left frontopolar cortices were shown in CA<or=2 group, with the left cingulate and right rostrodorsal prefrontal cortices showing prominent hypoperfusion. CA and PE (perseverative errors) scores significantly correlated with perfusion of the left posterior cingulate cortex. | | <https://pubmed.ncbi.nlm.nih.gov/16551505/> |
