## Supplementary Table 3 for "Agentic AI for automated hypothesis testing in Alzheimer’s disease and related dementias"

**Supplementary Table 3: Statistical analysis.** P-values were calculated using the Mann-Whitney U test (Wilcoxon rank-sum test), a two-sided non-parametric test that compares the distributions of counts between independent groups. For each hypothesis approval status category (NULL REJECTED, NULL NOT REJECTED, NOT TESTABLE), pairwise comparisons were performed across all three sample sizes (100, 1000, 10000). The test evaluates the null hypothesis that the two groups come from the same distribution.

| **Status** | Comparison | p-value |
| --- | --- | --- |
| **NULL REJECTED** | 100 vs 1000 | 0.012 |
| **NULL REJECTED** | 100 vs 10000 | 0.012 |
| **NULL REJECTED** | 1000 vs 10000 | 0.012 |
| **NULL NOT REJECTED** | 100 vs 1000 | 0.091 |
| **NULL NOT REJECTED** | 100 vs 10000 | 0.011 |
| **NULL NOT REJECTED** | 1000 vs 10000 | 0.093 |
| **NOT TESTABLE** | 100 vs 1000 | 0.011 |
| **NOT TESTABLE** | 100 vs 10000 | 0.011 |
| **NOT TESTABLE** | 1000 vs 10000 | 0.094 |
