## Supplementary Table 4 for "Agentic AI for automated hypothesis testing in Alzheimer’s disease and related dementias"

**Supplementary Table 4: Randomly generated hypotheses for negative control.** This table lists 100 hypotheses derived from general medical literature across diverse topics such as cardiovascular disease, infectious diseases, endocrinology, and oncology. These unrelated queries serve as a negative control to validate our agentic system’s specificity and robustness when applied to out-of-domain data in the NACC dataset.

**Hypothesis**

Higher body mass index is associated with increased risk of type 2 diabetes.

Regular exercise reduces cardiovascular disease mortality.

Smoking increases the risk of lung cancer by 20-fold.

Hypertension in midlife predicts later cardiovascular events.

Statin therapy reduces LDL cholesterol and cardiovascular risk.

Metformin improves insulin sensitivity in prediabetic patients.

Vitamin D deficiency is linked to increased fracture risk.

Chronic kidney disease progression is slowed by ACE inhibitors.

Obesity increases the risk of obstructive sleep apnea.

Regular mammography screening reduces breast cancer mortality.

Colonoscopy screening detects precancerous polyps effectively.

Antibiotic resistance is increasing in community-acquired pneumonia.

Influenza vaccination reduces hospitalization rates in elderly.

Hepatitis B vaccination prevents chronic liver disease.

Helicobacter pylori eradication reduces gastric cancer risk.

Proton pump inhibitors are effective for GERD symptom control.

Inflammatory bowel disease is associated with increased colorectal cancer risk.

Celiac disease screening improves outcomes in at-risk populations.

Liver cirrhosis increases the risk of hepatocellular carcinoma.

Pancreatic enzyme replacement improves malabsorption in chronic pancreatitis.

Beta-blockers reduce mortality in heart failure patients.

Aspirin reduces cardiovascular events in high-risk patients.

Warfarin prevents stroke in atrial fibrillation.

Coronary artery bypass grafting improves survival in multi-vessel disease.

Percutaneous coronary intervention is effective for acute MI.

Cardiac rehabilitation reduces readmission rates post-MI.

Hypertension control reduces stroke and heart attack risk.

Cholesterol-lowering diets reduce cardiovascular disease risk.

Omega-3 fatty acids may reduce triglyceride levels.

Sodium restriction lowers blood pressure in hypertensive patients.

Breastfeeding reduces the risk of childhood infections.

Childhood vaccination prevents serious infectious diseases.

Antibiotic stewardship reduces resistance development.

Hand hygiene compliance reduces hospital-acquired infections.

Isolation precautions prevent transmission of resistant organisms.

Surgical site infection rates decrease with proper protocols.

Central line bundles reduce bloodstream infections.

Ventilator-associated pneumonia decreases with elevation protocols.

Catheter-associated UTIs are reduced by early removal.

MRSA screening reduces transmission in hospitals.

Tuberculosis treatment adherence prevents drug resistance.

HIV antiretroviral therapy reduces transmission risk.

Hepatitis C treatment achieves high cure rates.

Malaria prophylaxis is effective in endemic regions.

Travel vaccinations prevent imported infectious diseases.

Seasonal flu vaccination reduces community spread.

Pneumococcal vaccination prevents invasive disease in elderly.

Shingles vaccination reduces postherpetic neuralgia risk.

HPV vaccination prevents cervical cancer development.

Meningococcal vaccination is effective in college settings.

Tetanus boosters prevent disease in wound injuries.

Diabetes screening identifies prediabetes early.

HbA1c monitoring guides diabetes management.

Insulin therapy improves glycemic control in type 1 diabetes.

GLP-1 agonists promote weight loss in type 2 diabetes.

SGLT2 inhibitors reduce heart failure hospitalizations.

Diabetic retinopathy screening prevents vision loss.

Diabetic nephropathy screening detects early kidney damage.

Diabetic foot care prevents amputations.

Thyroid function tests diagnose hypothyroidism accurately.

Levothyroxine replacement normalizes thyroid function.

Hyperthyroidism treatment prevents cardiac complications.

Calcium and vitamin D prevent osteoporosis fractures.

Bisphosphonates reduce fracture risk in osteoporosis.

Hormone replacement therapy relieves menopausal symptoms.

Testosterone replacement improves symptoms in hypogonadism.

Growth hormone therapy benefits children with deficiency.

Cortisol replacement is life-saving in adrenal insufficiency.

Parathyroid hormone monitoring guides calcium management.

Insulinoma resection cures hypoglycemia.

Pheochromocytoma removal prevents hypertensive crises.

Cushing syndrome treatment reverses metabolic abnormalities.

Addison disease requires lifelong steroid replacement.

Polycystic ovary syndrome increases diabetes risk.

Gestational diabetes screening improves pregnancy outcomes.

Preeclampsia monitoring prevents maternal complications.

Folic acid supplementation prevents neural tube defects.

Prenatal care reduces maternal and infant mortality.

Breast cancer screening detects early-stage disease.

Lumpectomy with radiation is equivalent to mastectomy for early cancer.

Chemotherapy improves survival in node-positive breast cancer.

Hormone therapy is effective for ER-positive breast cancer.

Prostate cancer screening detects disease early.

Active surveillance is appropriate for low-risk prostate cancer.

Radical prostatectomy cures localized prostate cancer.

Lung cancer screening reduces mortality in high-risk smokers.

Targeted therapy improves outcomes in EGFR-mutant lung cancer.

Immunotherapy extends survival in advanced lung cancer.

Colorectal cancer screening prevents cancer deaths.

Surgical resection cures early-stage colorectal cancer.

Adjuvant chemotherapy improves survival in stage 3 colon cancer.

Liver transplantation cures end-stage liver disease.

Kidney transplantation improves survival over dialysis.

Heart transplantation treats end-stage heart failure.

Lung transplantation improves quality of life in advanced disease.

Bone marrow transplantation cures certain hematologic malignancies.

Organ rejection is prevented by immunosuppressive drugs.

Graft-versus-host disease is managed with immunosuppression.

Post-transplant monitoring detects rejection early.

Living donor transplants have better outcomes than deceased donor.
